# We Are on the Verge of Breakthrough Cures for Type 1 Diabetes, but Who Are the 2 Million Americans Who Have It?

**DOI:** 10.1101/2024.07.24.24310877

**Authors:** Rebecca Smith, Samara Eisenberg, Aaron Turner-Pfifer, Jacqueline Le Grand, Sarah Pincus, Yousra Omer, Fei Wang, Bruce Pyenson

## Abstract

Two million Americans have Type 1 Diabetes. Innovative treatments have standardized insulin delivery and improved outcomes for patients, but patients’ access to such technologies depends on social determinants of health, including insurance coverage, proper diagnosis, and appropriate patient supports. Prior estimates of US prevalence, incidence, and patient characteristics have relied on data from select regions and younger ages and miss important determinants. By contrast, our research leveraged nationally representative administrative claims datasets to build a nuanced picture of the population with T1DM. Our work also supports future policy and research efforts with 2024, 2029, and 2033 projections of demographic and insurance coverage for people with T1DM.

## INTRODUCTION

The 2022 publication of global statistics for type 1 diabetes mellitus (T1DM) highlights the need for better statistics on the two million Americans with the condition.^1^ Recent technological advancements that customize T1DM therapies on a minute-by-minute patient basis produce improved outcomes for people with T1DM, and research points to potential cures.^2^ Better care will need to overcome stereotypes, such as T1DM’s legacy as “juvenile diabetes,” confusion with the more common Type 2 diabetes mellitus (T2DM), and incomplete demographic profiles based on limited registry information. Furthermore, US demographic and insurance coverage changes will affect population health efforts to reach people with T1DM. This paper fills gaps in US T1DM information with 2024, 2029, and 2033 projections of demographic and insurance coverage for people with T1DM.

The isolation of insulin in 1921 marked a transformative moment for what is now known as T1DM, turning it from a tragic and rapidly fatal childhood disease into a survivable chronic condition, although one with significant comorbidities and excess mortality.^3,4^ Recent progress has led to broad adoption of continuous glucose monitors (CGM), portable electronic devices that track and monitor glucose levels in real time, and wearable insulin pumps, which have reduced comorbidity burdens and increased survival rates for people with T1DM.^5^ Soon, we may have treatments to delay, prevent and even reverse the autoimmune processes that cause T1DM.^2^

Diabetes refers to several illnesses involving the body’s metabolization of sugar via insulin, a pancreatic enzyme. This paper focuses on T1DM, where the pancreas stops making insulin.^6^ T1DM becomes fatal quickly without regular injections of insulin.

Because T1DM was primarily diagnosed in children, it was originally called, “juvenile diabetes.” Type 2 diabetes, which is more prevalent and often associated with obesity, occurs when the pancreas produces insufficient insulin or ineffectively uses insulin for sugar metabolism. In both forms of diabetes, fluctuations in blood sugar (poor glycemic control) are associated with comorbidities. Although T1DM and T2DM are very different diseases, many information sources do not distinguish between the conditions; patient diagnoses may be miscoded, and patients’ forms of diabetes may not be obvious from their treatments.^7,8^

About 2 million Americans have T1DM—much lower than the nearly 30 million with T2DM.^9^ Diabetes is the 8^th^ leading cause of death in the US with about 9% of deaths coming from T1DM.^10^ T1DM is predominantly diagnosed in children and young adults but similarly across sexes.^6^ Patients with T1DM have mortality rates 3-18 times higher than standard.^11^ Innovative treatments and technologies have helped standardize insulin delivery and improve outcomes for patients. A variety of automated devices are in use, such as continuous glucose monitors (CGMs) and insulin pumps, but patients’ access to such devices depends on insurance coverage, proper diagnosis, and appropriate patient supports, all of which are affected by social determinants of health.^12^

Our use of real-world data, which includes some key socioeconomic drivers, provides important public health information about the 2 million Americans with T1DM. Historically, estimates of T1DM prevalence and incidence have come from epidemiological, clinic-based, population-based, prospective birth, case cohort, and cross-sectional studies.^13^ By contrast, we used several large, nationwide payer-based administrative datasets combined with estimates of incidence changes and US demographic projections by region and ethnicity to produce 5- and 10-year forecasts. We found that the number of T1DM patients is higher than other estimates, with important regional and socioeconomic differences, some of which are expected to widen over time. Finally, we replicated others’ findings of significant T1DM incidence among older adults, including those covered by Medicare.^14^

We hope forecasting the demographic details of America’s population with T1DM will help public health, payer, and advocates’ efforts to spread best practices and, optimistically, future cures.

## DATA AND METHODS

This study utilized real-world administrative claims data (which insurers and others collect when they process bills from healthcare providers during their payments for covered services, devices, and drugs,) to create estimates of the US T1DM population in 2019, segmented by type of insurance coverage (e.g., commercial, Medicaid, Medicare, uninsured, etc.).

### Claims Analysis

We analyzed 2018 to 2020 data from several large administrative databases (Appendix Exhibit A1).^15–17^ Patients were required to have at least 1 month of enrollment between January 2018 and December 2020, except for the Medicare Advantage (MA) population for which data were available through December 2019.

Insulin-using patients were identified using 3+ distinct claims for insulin, insulin pumps, or insulin-related supplies on separate dates, at least 30 to 120 days apart (Appendix Exhibit A2-A3). Patients with T1DM were identified from among the insulin-using population using a claims-based algorithm (Appendix Exhibit A2-A4). Patients’ index dates were set to the date of their first claim with a T1DM diagnosis. Patients with evidence of drug combinations specific to T2DM at any time during the study were excluded (Appendix Exhibit A5).^8^ Additionally, patients with evidence of only long-acting insulin prescriptions (Appendix Exhibit A2-A3) were excluded; patients using pre-mixed insulin formulations including a short- or intermediate-acting insulin (Appendix Exhibit A2-A3) were included.

Patients were flagged as newly diagnosed with T1DM (“incident”) if there was no evidence of a T1DM diagnosis (Appendix Exhibit A4) or use of insulin, insulin-related durable medical equipment (DME), or an insulin pump (Appendix Exhibit A2-A3) within 6 months of their index date. This identification approach is similar to other published and validated algorithms.^8,18^

Patient counts were compiled for each dataset based on insurance coverage (Commercial (COM), Medicaid (MCD), Medicare fee-for-service (FFS), MA), age, sex, geographic region, and metropolitan statistical area (MSA) status. For the Medicare populations, additional variables were captured, including race/ethnicity and dual-Medicare-Medicaid eligibility status, the latter indicating low-income beneficiaries.

Patients were grouped into five-year age bands, with wider bands for the youngest and oldest ages. We used four geographic regions (Appendix Exhibit A6) and split residence into MSA (urban) or not (rural). Sex was captured from enrollment data, and race/ethnicity categories tabulated included African American, Hispanic, non-Hispanic white, and all other races (available only for Medicare).

### Extrapolation of Claims Data to National Estimates

Prevalence rates were determined for each of the coverages analyzed and extrapolated to national counts for 2019. (Appendix Exhibits A1 and A7).^19–24^ In addition to the four coverage types directly examined via claims, we developed prevalence rates for Veterans Affairs (VA), Uninsured, and Other Medicare (individuals with either only Medicare Part A or only Part B). The commercial prevalence rate was used for the VA population, the Managed MCD prevalence rate for all MCD populations and the Uninsured, and the FFS prevalence rate for the Other Medicare population.

For modeling, claims-derived patient counts were used to calculate agent weights for each combination of demographic factors within each insurance coverage.

T1DM incidence rates were computed across coverage-specific age, sex, demographic, and geographic cohorts. For rates of T1DM device use, patients in each cohort were categorized based on their usage of CGMs, insulin pumps, both CGMs and pumps, or neither device. T1DM age distributions, incidence rates by age, and device use rates by age were smoothed by fitting curves to initially developed rates derived from the data.

### 10-year Projections

Events within a given year, such as device uptake or death, were modeled in a probabilistic manner. We developed mortality loads for the T1DM population relative to standard population mortality separately for ages 0-64 and for 65+ using FFS data. We examined raw counts and compared actual deaths to those implied by standard mortality tables from CDC WONDER (Appendix Exhibits A8-A9).^25^

We note that our method of using Medicare-based mortality loads for commercial or Medicaid T1DM patients may overstate expected deaths because under-65 Medicare T1DM patients may suffer from disabling conditions such as end-stage renal disease.

The number of new cases of T1DM in the model were calibrated to produce a baseline, steady state model by age, which considered mortality and age progression. The baseline model maintained consistency in demographics for the T1DM population over the 10-year projection. Population growth was then incorporated into that model using US population growth projections^26^ (Appendix Exhibits A10-A11) and T1DM diabetes incidence growth from the Global Burden of Disease Study 2017.^27^

Mortality for patients newly starting on CGMs or insulin pump devices reflected the mortality-reducing impacts of these technologies.^28–31^ For patients using both devices, excess T1DM mortality over standard mortality was reduced by 50% (Appendix Exhibit A12). This improvement was applied in the first year of new device use and all subsequent years. For patients exclusively using CGMs, a 40% reduction was applied. For those exclusively using pumps, a 10% mortality reduction was applied. The model also assumed 85% of the population would use devices by Model Year 3, compared to approximately 78% in 2019.

## RESULTS

### 2024 Baseline

We estimated 2.07 million T1DM patients nationally across all insurance coverages in our 2024 baseline model year – 1.79 million adults (20+) and 0.28 million children (Exhibit 1 Table 1). This represents a US T1DM prevalence rate of 617 per 100,000 (Exhibit 2 Figure 1) with an average age of 47. The majority of patients (68%) were classified as Non-Hispanic White, and the largest proportion were covered by COM (47%), followed by Medicare (FFS, MA, and Other Medicare populations totaling 29%) and Medicaid (15%) insurance coverage (Exhibit 4 Table 2). We observed 78% of patients with CGMs and/or insulin pump devices.

**EXHIBIT 1 (Table 1).** United States Model Years 1, 5, and 10 - T1DM patient count projections for each demographic group and estimated device use.

| T1DM Patients | Model Years |  |  | Cumulative % Change |  |  |
| --- | --- | --- | --- | --- | --- | --- |
| Population Group | 1 | 5 | 10 | Years 1-5 | Years 5-10 | Years 1-10 |
| **Total (N)** |  |  |  |  |  |  |
| **Total Population** | 2,072,557 | 2,148,483 | 2,289,197 | 3.7% | 6.5% | 10.5% |
| **By Payer (N)** |  |  |  |  |  |  |
| **Commercial** | 973,020 | 983,365 | 1,008,562 | 1.1% | 2.6% | 3.7% |
| **Medicaid** | 311,021 | 345,386 | 404,094 | 11.0% | 17.0% | 29.9% |
| **Medicare Fee-for-Service** | 362,655 | 356,818 | 359,743 | -1.6% | 0.8% | -0.8% |
| **Medicare Advantage** | 210,443 | 230,923 | 262,413 | 9.7% | 13.6% | 24.7% |
| **Veterans Affairs** | 23,883 | 24,423 | 26,494 | 2.3% | 8.5% | 10.9% |
| **Uninsured** | 157,063 | 171,856 | 189,568 | 9.4% | 10.3% | 20.7% |
| **Medicare Part A Only or Part B only** | 34,472 | 35,713 | 38,322 | 3.6% | 7.3% | 11.2% |
| **By Sex (N)** |  |  |  |  |  |  |
| **Female** | 1,031,497 | 1,075,807 | 1,152,395 | 4.3% | 7.1% | 11.7% |
| **Male** | 1,041,060 | 1,072,676 | 1,136,802 | 3.0% | 6.0% | 9.2% |
| **By Age Band (N)** |  |  |  |  |  |  |
| **<10** | 56,280 | 60,787 | 77,708 | 8.0% | 27.8% | 38.1% |
| **10-14** | 96,241 | 99,259 | 108,754 | 3.1% | 9.6% | 13.0% |
| **15-19** | 129,387 | 131,059 | 135,831 | 1.3% | 3.6% | 5.0% |
| **20-24** | 140,183 | 140,724 | 142,202 | 0.4% | 1.1% | 1.4% |
| **25-29** | 144,392 | 144,962 | 146,677 | 0.4% | 1.2% | 1.6% |
| **30-34** | 148,645 | 149,347 | 151,345 | 0.5% | 1.3% | 1.8% |
| **35-39** | 152,902 | 153,776 | 156,162 | 0.6% | 1.6% | 2.1% |
| **40-44** | 157,155 | 158,236 | 161,089 | 0.7% | 1.8% | 2.5% |
| **45-49** | 161,402 | 162,811 | 166,477 | 0.9% | 2.3% | 3.1% |
| **50-54** | 165,348 | 167,217 | 171,791 | 1.1% | 2.7% | 3.9% |
| **55-59** | 162,579 | 164,825 | 170,394 | 1.4% | 3.4% | 4.8% |
| **60-64** | 159,402 | 163,020 | 171,854 | 2.3% | 5.4% | 7.8% |
| **65+** | 398,640 | 452,461 | 528,914 | 13.5% | 16.9% | 32.7% |
| **By Region (N)** |  |  |  |  |  |  |
| **Midwest** | 529,932 | 548,840 | 581,709 | 3.6% | 6.0% | 9.8% |
| **Northeast** | 400,337 | 410,041 | 432,965 | 2.4% | 5.6% | 8.2% |
| **South** | 749,807 | 772,540 | 814,895 | 3.0% | 5.5% | 8.7% |
| **West** | 392,481 | 417,063 | 459,627 | 6.3% | 10.2% | 17.1% |
| **By Race (N)** |  |  |  |  |  |  |
| **African-American** | 327,663 | 344,542 | 370,007 | 5.2% | 7.4% | 12.9% |
| **Hispanic** | 268,984 | 286,829 | 317,191 | 6.6% | 10.6% | 17.9% |
| **Non-Hispanic White** | 1,403,788 | 1,438,405 | 1,513,058 | 2.5% | 5.2% | 7.8% |
| **All Other Race** | 72,122 | 78,707 | 88,940 | 9.1% | 13.0% | 23.3% |
| **By Dual Status (N)** |  |  |  |  |  |  |
| **Dual** | 160,448 | 170,590 | 187,326 | 6.3% | 9.8% | 16.8% |
| **Non-Dual** | 447,122 | 452,864 | 473,152 | 1.3% | 4.5% | 5.8% |
| **By MSA Status** |  |  |  |  |  |  |
| **MSA** | 1,880,100 | 1,954,650 | 2,091,173 | 4.0% | 7.0% | 11.2% |
| **Non-MSA** | 192,457 | 193,833 | 198,024 | 0.7% | 2.2% | 2.9% |
| **Average Age** | 47.2 | 48.1 | 48.9 | 1.8% | 1.7% | 3.5% |

Source: Authors' analysis based on Milliman's Consolidated Health Cost Guidelines Sources Database (CHSD), IBM MarketScan®, and the Centers for Medicare and Medicaid Services (CMS) 100% Research Identifiable Files (RIF). Notes: This exhibit shows projected T1DM patient counts under each insurance type, gender, age band, region, race, dual status (for Medicare patients), and metropolitan statistical area (MSA) status and in total. The three columns on the far right represent the percent change over the specified time interval. Growth in each demographic group tends to speed up in the later model years.

**EXHIBIT 2 (Figure 1).**
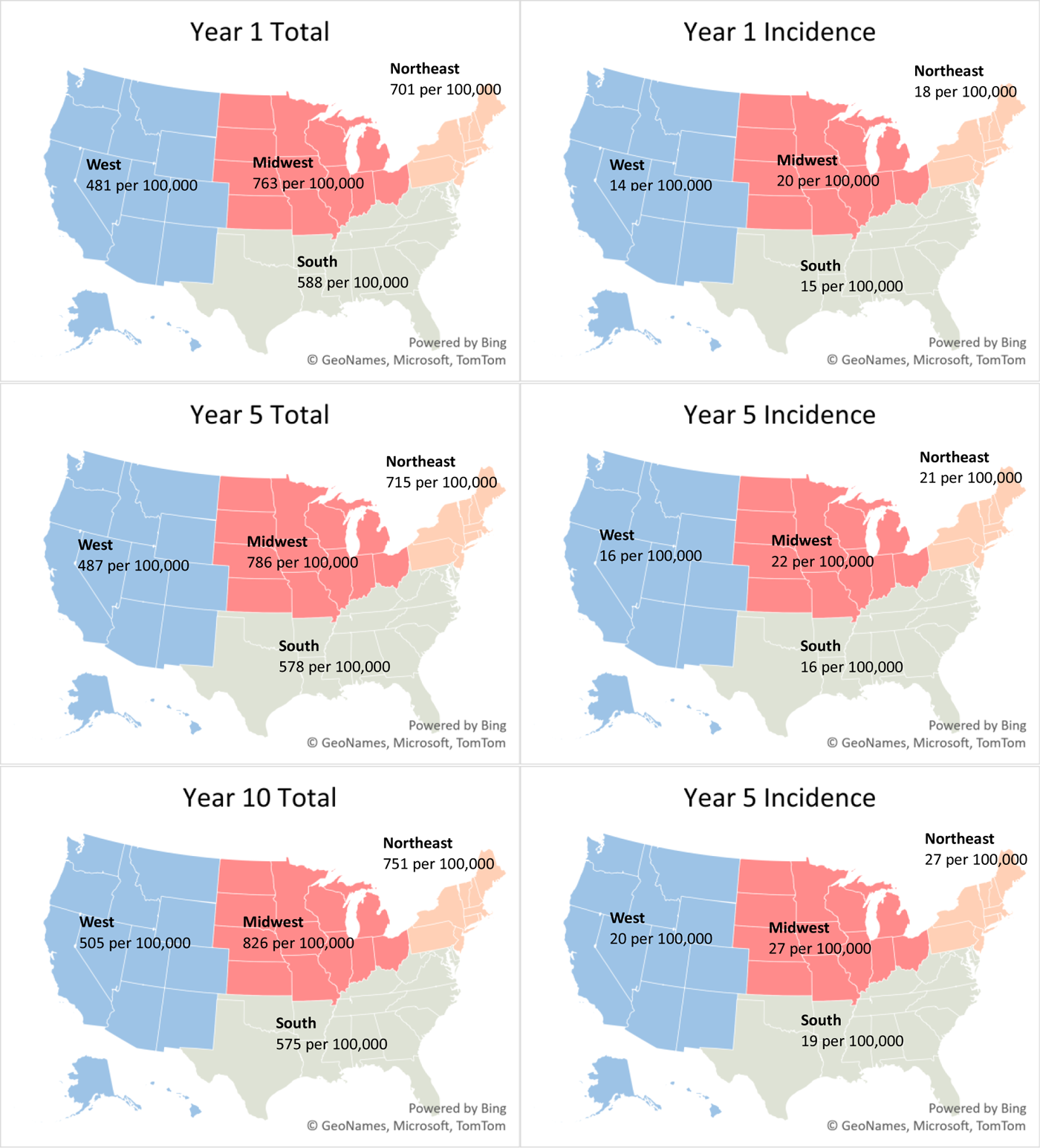
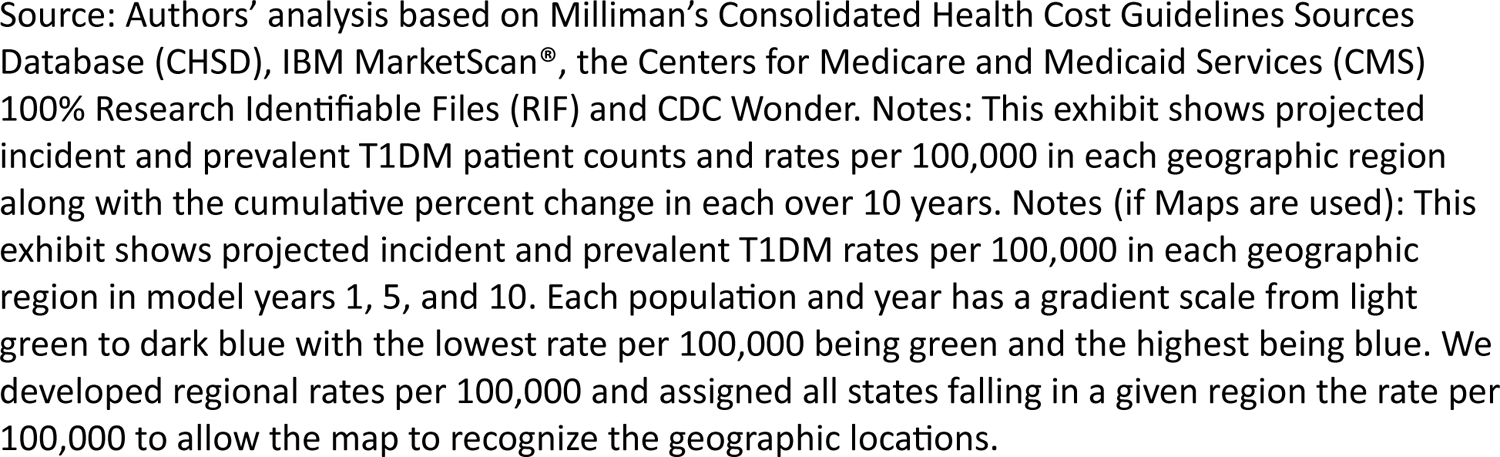
Variation of incident and prevalent T1DM by geographic region in model years 1, 5, and 10

Incident patients were approximately 2.6% of the total patient population, an incidence rate of 0.016%. About 14% of the incident population over the projection period were ≥65 years old (Exhibit 3 Figure 2). Regionally, the Midwest and Northeast exhibited the highest baseline incidence rates at 20 and 18 per 100,000, respectively (Exhibit 2 Figure 1). These rates are 25-35% higher than observed incidence rates in the South and the West (15 and 14 per 100,000, respectively). About 47% of incident patients were covered under commercial insurance followed by Medicare (21%) and Medicaid (20%) (Exhibit 4 Table 2).

**EXHIBIT 3 (Figure 2).**
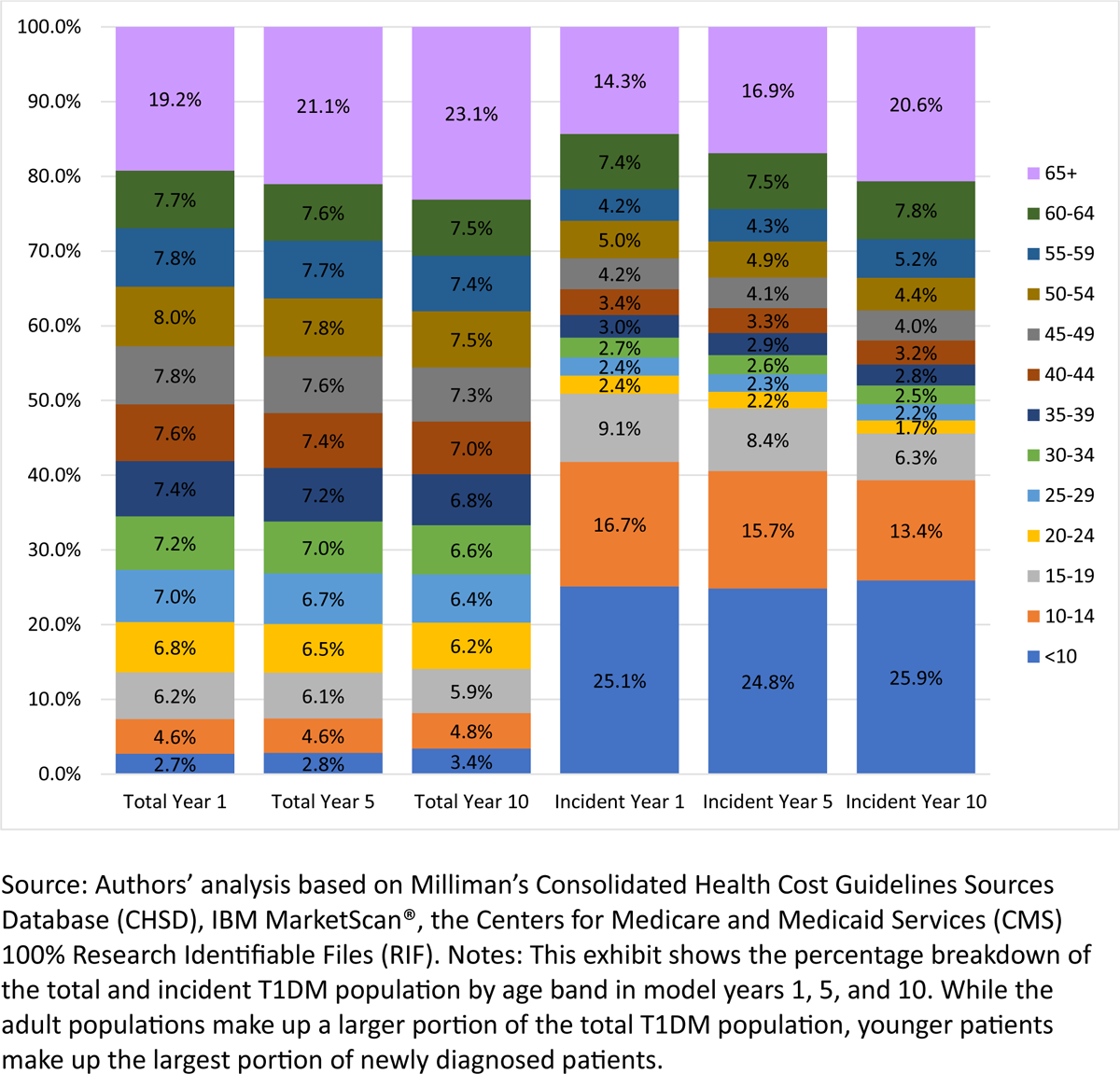
Distribution of total T1DM and incident T1DM populations by age band in model years 1, 5, and 10

**EXHIBIT 4 Table 2.** United States Model Year 1 T1DM patient count projections and distribution by age and sex within each insurance market.

| Sex | Age Band | Total | COM | MCD | FFS | MA | VA | UNI | Other MCR |
| --- | --- | --- | --- | --- | --- | --- | --- | --- | --- |
| **Male Total** | **All Ages** | 1,041,060 | 520,423 | 150,344 | 170,579 | 94,792 | 12,774 | 75,920 | 16,228 |
| **Male** | **<10** | 29,460<br>(2.8%) | 14,777<br>(2.8%) | 9,529<br>(6.3%) | 0<br>(0.0%) | 0<br>(0.0%) | 365<br>(2.9%) | 4,789<br>(6.3%) | 0<br>(0.0%) |
| **Male** | **10-14** | 48,713<br>(4.7%) | 26,639<br>(5.1%) | 14,239<br>(9.5%) | 0<br>(0.0%) | 0<br>(0.0%) | 655<br>(5.1%) | 7,180<br>(9.5%) | 0<br>(0.0%) |
| **Male** | **15-19** | 69,069<br>(6.6%) | 40,238<br>(7.7%) | 18,501<br>(12.3%) | 0<br>(0.0%) | 0<br>(0.0%) | 988<br>(7.7%) | 9,342<br>(12.3%) | 0<br>(0.0%) |
| **Male** | **20-24** | 72,562<br>(7.0%) | 50,412<br>(9.7%) | 13,685<br>(9.1%) | 284<br>(0.2%) | 2<br>(0.0%) | 1,237<br>(9.7%) | 6,915<br>(9.1%) | 27<br>(0.2%) |
| **Male** | **25-29** | 71,822<br>(6.9%) | 47,610<br>(9.1%) | 14,484<br>(9.6%) | 1,075<br>(0.6%) | 64<br>(0.1%) | 1,168<br>(9.1%) | 7,319<br>(9.6%) | 102<br>(0.6%) |
| **Male** | **30-34** | 75,361<br>(7.2%) | 47,265<br>(9.1%) | 15,356<br>(10.2%) | 3,095<br>(1.8%) | 435<br>(0.5%) | 1,160<br>(9.1%) | 7,759<br>(10.2%) | 292<br>(1.8%) |
| **Male** | **35-39** | 78,344<br>(7.5%) | 48,249<br>(9.3%) | 14,230<br>(9.5%) | 5,691<br>(3.3%) | 1,263<br>(1.3%) | 1,184<br>(9.3%) | 7,190<br>(9.5%) | 537<br>(3.3%) |
| **Male** | **40-44** | 82,206<br>(7.9%) | 49,947<br>(9.6%) | 13,392<br>(8.9%) | 7,759<br>(4.5%) | 2,383<br>(2.5%) | 1,226<br>(9.6%) | 6,767<br>(8.9%) | 732<br>(4.5%) |
| **Male** | **45-49** | 85,192<br>(8.2%) | 51,497<br>(9.9%) | 11,736<br>(7.8%) | 9,913<br>(5.8%) | 3,916<br>(4.1%) | 1,264<br>(9.9%) | 5,930<br>(7.8%) | 935<br>(5.8%) |
| **Male** | **50-54** | 86,850<br>(8.3%) | 51,399<br>(9.9%) | 10,118<br>(6.7%) | 11,874<br>(7.0%) | 5,965<br>(6.3%) | 1,261<br>(9.9%) | 5,112<br>(6.7%) | 1,120<br>(6.9%) |
| **Male** | **55-59** | 83,780<br>(8.0%) | 47,310<br>(9.1%) | 8,763<br>(5.8%) | 12,948<br>(7.6%) | 7,951<br>(8.4%) | 1,161<br>(9.1%) | 4,428<br>(5.8%) | 1,219<br>(7.5%) |
| **Male** | **60-64** | 80,550<br>(7.7%) | 45,079<br>(8.7%) | 6,312<br>(4.2%) | 13,648<br>(8.0%) | 9,930<br>(10.5%) | 1,106<br>(8.7%) | 3,188<br>(4.2%) | 1,287<br>(7.9%) |
| **Male** | **65+** | 177,152<br>(17.0%) | 0<br>(0.0%) | 0<br>(0.0%) | 104,290<br>(61.1%) | 62,883<br>(66.3%) | 0<br>(0.0%) | 0<br>(0.0%) | 9,978<br>(61.5%) |
| **Female Total** | **All Ages** | 1,031,497 | 452,597 | 160,677 | 192,076 | 115,651 | 11,109 | 81,143 | 18,244 |
| **Female** | **<10** | 26,820<br>(2.6%) | 13,240<br>(2.9%) | 8,821<br>(5.5%) | 0<br>(0.0%) | 0<br>(0.0%) | 327<br>(2.9%) | 4,433<br>(5.5%) | 0<br>(0.0%) |
| **Female** | **10-14** | 47,529<br>(4.6%) | 24,593<br>(5.4%) | 14,845<br>(9.2%) | 0<br>(0.0%) | 0<br>(0.0%) | 605<br>(5.4%) | 7,486<br>(9.2%) | 0<br>(0.0%) |
| **Female** | **15-19** | 60,318<br>(5.8%) | 34,872<br>(7.7%) | 16,339<br>(10.2%) | 0<br>(0.0%) | 0<br>(0.0%) | 856<br>(7.7%) | 8,251<br>(10.2%) | 0<br>(0.0%) |
| **Female** | **20-24** | 67,621<br>(6.6%) | 42,569<br>(9.4%) | 15,852<br>(9.9%) | 132<br>(0.1%) | 1<br>(0.0%) | 1,044<br>(9.4%) | 8,010<br>(9.9%) | 12<br>(0.1%) |
| **Female** | **25-29** | 72,570<br>(7.0%) | 43,374<br>(9.6%) | 18,052<br>(11.2%) | 843<br>(0.4%) | 36<br>(0.0%) | 1,064<br>(9.6%) | 9,122<br>(11.2%) | 80<br>(0.4%) |
| **Female** | **30-34** | 73,284<br>(7.1%) | 41,256<br>(9.1%) | 18,076<br>(11.2%) | 3,100<br>(1.6%) | 413<br>(0.4%) | 1,012<br>(9.1%) | 9,134<br>(11.3%) | 293<br>(1.6%) |
| **Female** | **35-39** | 74,558<br>(7.2%) | 42,739<br>(9.4%) | 15,700<br>(9.8%) | 5,129<br>(2.7%) | 1,525<br>(1.3%) | 1,049<br>(9.4%) | 7,933<br>(9.8%) | 484<br>(2.7%) |
| **Female** | **40-44** | 74,949<br>(7.3%) | 41,913<br>(9.3%) | 14,142<br>(8.8%) | 7,335<br>(3.8%) | 2,694<br>(2.3%) | 1,028<br>(9.3%) | 7,146<br>(8.8%) | 691<br>(3.8%) |
| **Female** | **45-49** | 76,211<br>(7.4%) | 43,223<br>(9.5%) | 11,684<br>(7.3%) | 9,117<br>(4.7%) | 4,363<br>(3.8%) | 1,061<br>(9.5%) | 5,904<br>(7.3%) | 859<br>(4.7%) |
| **Female** | **50-54** | 78,497<br>(7.6%) | 44,027<br>(9.7%) | 10,164<br>(6.3%) | 10,865<br>(5.7%) | 6,201<br>(5.4%) | 1,080<br>(9.7%) | 5,135<br>(6.3%) | 1,024<br>(5.6%) |
| **Female** | **55-59** | 78,799<br>(7.6%) | 41,505<br>(9.2%) | 9,323<br>(5.8%) | 12,547<br>(6.5%) | 8,513<br>(7.4%) | 1,018<br>(9.2%) | 4,711<br>(5.8%) | 1,181<br>(6.5%) |
| **Female** | **60-64** | 78,852<br>(7.6%) | 39,286<br>(8.7%) | 7,679<br>(4.8%) | 14,287<br>(7.4%) | 11,410<br>(9.9%) | 964<br>(8.7%) | 3,879<br>(4.8%) | 1,346<br>(7.4%) |
| **Female** | **65+** | 221,488<br>(21.5%) | 0<br>(0.0%) | 0<br>(0.0%) | 128,720<br>(67.0%) | 80,494<br>(69.6%) | 0<br>(0.0%) | 0<br>(0.0%) | 12,274<br>(67.3%) |

Source: Authors' analysis based on Milliman's Consolidated Health Cost Guidelines Sources Database (CHSD), IBM MarketScan®, the Centers for Medicare and Medicaid Services (CMS) 100% Research Identifiable Files (RIF). Notes: This exhibit shows the count and percentage age band breakdown of T1DM patients within each sex for each insurance market and in total. COM represents Commercial, MCD represents Medicaid, FFS represents Medicare Fee-for-Service, MA represents Medicare Advantage, VA represents Veterans Affairs, UNI represents Uninsured, and Other MCR represents the Other Medicare population.

Claims-derived mortality among patients with T1DM were roughly three times those of the overall population. However, after incorporating mortality improvement associated with device use, T1DM mortality decreased to around twice that of the overall population.

### 10-Year Projection

By 2033, we project the US population with T1DM will grow by about 10%, reaching approximately 2.29 million patients from 2.27 in 2024. This growth is attributed to a nearly 50% increase in the number of incident patients over the next decade coupled with improved survival from use of devices. Over the decade, the average age of patients is expected to increase from 47 to 49, and the number of patients over age 65 is projected to increase by 33% (Exhibit 1 Table 1). The over 65 growth is primarily influenced by the aging of the US population rather than by increases in T1DM incidence among older Americans.

The largest growth in T1DM incidence is expected in the Northeast. However, due to expected US population growth patterns, T1DM prevalence in the West will increase most (17%).

The racial and ethnic composition of the T1DM population is expected to align with overall US trends. The Hispanic population with T1DM is projected to grow by 18% and the African American population with T1DM by nearly 13%. Although Non-Hispanic White patients will continue to constitute the majority of the US T1DM population, that share is projected to decrease from 68% to 66%, and 10-year growth among Non-Hispanic White patients with T1DM will be less than 10%, the lowest among all racial and ethnic groups modeled (Exhibit 1 Table 1).

MA and MCD are expected to undergo the most substantial increases in T1DM growth over the next decade, with patient populations growing by 25% and 30% respectively. Projected growth in T1DM among MA-covered patients reflects ongoing, known shifts in Medicare coverage from FFS to MA (Exhibit 1 Table 1).

## DISCUSSION

Our analysis produces somewhat different results than other studies. The most detailed estimates of T1DM among US youth derive from the SEARCH for Diabetes in Youth studies, which examined populations in 10 states.^32^ Northeast states were not included in SEARCH. However, our nationwide approach found the Northeast has the highest T1DM incidence. This is one reason our estimated T1DM population is larger than others’. Large regional variation in incidence has been observed in international studies. For example, a 2020 study by Mobasseri et al. found America had the highest incidence compared to Asia, Africa, and Europe.^33, 31^ Another study among children aged 0-4 found western European regions had the highest incidence compared to other world regions in this age group.^34^ We combined US regional incidence differences with population forecasts that include regional and socioeconomic factors. The results show important differences across regions, payers, and ethnic groups. We found T1DM prevalence tends to vary by income, which is consistent with several international studies that found more developed countries observe higher incidence and prevalence than less developed countries.^1,33^ It is possible that higher patient income is associated with more accurate coding in clinical or administrative data.

Much existing literature on T1DM prevalence and incidence is based on epidemiological, clinic-based, population-based, prospective birth, case cohort, and cross-sectional studies.^13^ These studies may have limitations such as inadequate representation for the full population, insufficient detail on potential additional contributing factors, limited sample sizes, challenges in control selection, and bias in self-reported data, particularly in survey-based methodologies. By contrast, our study used real-world data from claims to determine T1DM estimates. Advantages of claims studies include data quality and consistency, clinical validity, ability to link demographic variables, and broad data availability, but we recognize other limitations.^35^ For example, we identified individuals with T1DM using various fields including diagnosis, procedure, and drug codes. These fields may be underreported or misreported. Payment for drugs or devices does not guarantee actual patient use. Indeed, our estimates of device use were based on claims for these devices, but we did not assess adherence, so our estimates may overstate actual, ongoing utilization.

Distinguishing between T1DM and T2DM poses challenges both clinically and epidemiologically. T1DM is the less common condition, and many T1DM cases may be coded as T2DM. Adult onset T1DM may be especially subject to miscoding due to the incorrect perception that adult cases are rare.

CGMs and insulin pumps have become the standard of care under US and other clinical guidelines^36,37^, and use is increasing.^31^ Their adoption is associated with decreased complications including hypoglycemia, diabetic ketoacidosis, and diabetes-related emergency visits.^31^ Our model incorporated projected increases in uptake for devices over time to reflect their clinical value and recent trends. As device use increases, we anticipate a reduction in complications and, consequently, projected deaths. However, each payer may implement specific coverage criteria or requirements, which can limit or delay access.

Commercially insured patients comprised the largest portion of our model, followed by Medicare and Medicaid. According to the Medicare Local Coverage Determination (LCD) which determines the “reasonable and necessary” criteria for CGM coverage^38^, CGMs are only covered when the following criteria are met:

1. The beneficiary has diabetes based on ICD-10 codes
2. The beneficiary is administered insulin 3+ times daily
3. The beneficiary’s treatment requires regular adjustment
4. Within six months prior to ordering the CGM, the beneficiary must have an inpatient visit with their treating practitioner confirming (1-3) are met
5. The beneficiary must have an in-person follow-up every six months to assess adherence.

FFS patients who meet the above criteria are eligible for coverage of their devices with few restrictions based on brand or cost. But access to therapies among commercial insurance plans can differ significantly. Commercial insurance plans may categorize insulin on different formulary tiers, resulting in varying coverage and out-of-pocket costs. Additionally, devices may be subject to insurer approval based on medical necessity criteria.

The mortality loads developed from claims data for patients with T1DM were nearly three times those of the general population, consistent with published findings.^39^ T1DM death counts produced by our model were also consistent with previously published mortality studies.^1^ Our modeling explored the expected life years gained based on mortality improvement from device use over the 10-year projection. We estimate this improvement will result in nearly 360,000 additional life years, a 2% increase, compared to the baseline scenario. While literature suggests that device use may improve risk factors for comorbidities, there is limited literature available on the quantitative impact that devices may have on mortality.

A recent study published identified a global increase of 60-107% in T1DM prevalence from 2021 to 2040.^40^ In contrast, our study suggests a 10% US increase from 2024 to 2033. Our study was limited to the US, a more developed country. Prevalence of T1DM is higher in more developed countries, but growth in prevalence tends to be higher in less developed countries.^1^ As technology for diagnosing and treating T1DM becomes more widely available, we expect developing countries to show greater T1DM growth.

Our study found lower incidence rates in middle age compared to children and young adults, consistent with prior research.^40^ However, when viewed as a whole, we observed substantial incidence across middle and older age brackets. As new-onset T1DM is more commonly misdiagnosed as T2DM in adults than in children^41^, our findings suggest there are public health implications to missing older individuals when considering how best to identify, treat, and support patients with this disease.

Our Year 1 (2024) T1DM population size is based on observed rates from 2019 data. Trending the data to 2024 introduces uncertainties, partly because of the disruption caused by the COVID-19 pandemic. Indeed, all population forecasts involve uncertainty because of economic, demographic, and clinical changes. By example, there have likely been changes in patient outcomes and mortality since 2019 due to the entrance and increased use of closed loop insulin delivery systems which combine CGM and pump.

Race and patients’ dates of death were available only in the FFS data set. The race field is self-reported, introducing potential inaccuracies. Mortality loads for all ages and payers were developed using FFS data, and while these loads are akin to those reported in literature, they may not be appropriate for other coverages. Of course, models are simplifications of reality, and assumptions applied for modeling purposes will likely differ from future experience.

Finally, the commercial databases we used are comprised of claims primarily from patients covered by large, self-insured employer-sponsored health plans with relatively rich benefits compared to Medicare or Medicaid. These data are recorded for the purpose of payment, not clinical intent, and thus are imperfect when clinical assumptions are applied. Additionally, MA data was available only through 2019, so our initial T1DM identification period was shorter for that market. Finally, we could not access data for certain populations, such as the Medicaid FFS, VA, and Uninsured populations, so we used proxies. Analyses using different years, data sources, methodologies may produce different results.

## CONCLUSION

T1DM impacts two million people in the US. Despite advances in technology and care management, these patients face high comorbidity and mortality risks, and T1DM prevalence continues to grow. But today also sees rapid evolution in our understanding and ability to treat T1DM. Clinicians can now screen for future risk of developing T1DM via blood test; multiple human clinical trials are underway for cell therapies that could end T1DM patients’ reliance on external insulin, and in 2022 the FDA approved the first disease-modifying therapy delaying T1DM onset. Given this rapidly changing landscape, data about the T1DM community is essential to ensure informed decisions by key stakeholders. This study represents a step toward a detailed understanding of the future composition of the T1DM population.

## Data Availability

All data produced in the present study are available upon reasonable request to the authors

---

Figure data

Data for rates per 100,000 by region in model years 1, 5, and 10.

| **T1DM Patients** | **Model Years** |  |  | **Cumulative**<br>**% Change** |
| --- | --- | --- | --- | --- |
| **Population Group** | **1** | **5** | **10** | **Years**<br>**1-10** |
| **Total** |  |  |  |  |
| **Total T1DM Population** | 2,072,557 | 2,148,483 | 2,289,197 | 10.5% |
| **Incident T1DM Population** | 54,454 | 63,605 | 79,914 | 46.8% |
| **T1DM Rate per 100,000** | 617.19 | 619.78 | 634.64 | 2.8% |
| **T1DM Incidence Rate per 100,000** | 16.22 | 18.35 | 22.15 | 36.6% |
| **T1DM Population Counts** |  |  |  |  |
| **Midwest** | 529,932 | 548,840 | 581,709 | 9.8% |
| **Northeast** | 400,337 | 410,041 | 432,965 | 8.2% |
| **South** | 749,807 | 772,540 | 814,895 | 8.7% |
| **West** | 392,481 | 417,063 | 459,627 | 17.1% |
| **Incident T1DM Population Counts** |  |  |  |  |
| **Midwest** | 13,642 | 15,598 | 19,198 | 40.7% |
| **Northeast** | 10,333 | 11,999 | 15,524 | 50.2% |
| **South** | 18,927 | 21,995 | 27,366 | 44.6% |
| **West** | 11,552 | 14,013 | 17,826 | 54.3% |
| **T1DM Rate per 100,000** |  |  |  |  |
| **Midwest** | 762.98 | 785.51 | 826.38 | 8.3% |
| **Northeast** | 700.68 | 714.99 | 751.45 | 7.2% |
| **South** | 587.76 | 578.11 | 575.43 | -2.1% |
| **West** | 480.73 | 486.70 | 504.87 | 5.0% |
| **T1DM Incidence Rate per 100,000** |  |  |  |  |
| **Midwest** | 19.64 | 22.32 | 27.27 | 38.8% |
| **Northeast** | 18.08 | 20.92 | 26.94 | 49.0% |
| **South** | 14.84 | 16.46 | 19.32 | 30.2% |
| **West** | 14.15 | 16.35 | 19.58 | 38.4% |

| Category | Region | State | Year | Rate per 100,000 |
| --- | --- | --- | --- | --- |
| Total | West | Alaska | 1 | 480.73 |
| Total | South | Alabama | 1 | 587.76 |
| Total | South | Arkansas | 1 | 587.76 |
| Total | West | Arizona | 1 | 480.73 |
| Total | West | California | 1 | 480.73 |
| Total | West | Colorado | 1 | 480.73 |
| Total | Northeast | Connecticut | 1 | 700.68 |
| Total | South | District of Columbia | 1 | 587.76 |
| Total | South | Delaware | 1 | 587.76 |
| Total | South | Florida | 1 | 587.76 |
| Total | South | Georgia | 1 | 587.76 |
| Total | West | Hawaii | 1 | 480.73 |
| Total | Midwest | Iowa | 1 | 762.98 |
| Total | West | Idaho | 1 | 480.73 |
| Total | Midwest | Illinois | 1 | 762.98 |
| Total | Midwest | Indiana | 1 | 762.98 |
| Total | Midwest | Kansas | 1 | 762.98 |
| Total | South | Kentucky | 1 | 587.76 |
| Total | South | Louisiana | 1 | 587.76 |
| Total | Northeast | Massachusetts | 1 | 700.68 |
| Total | South | Maryland | 1 | 587.76 |
| Total | Northeast | Maine | 1 | 700.68 |
| Total | Midwest | Michigan | 1 | 762.98 |
| Total | Midwest | Minnesota | 1 | 762.98 |
| Total | Midwest | Missouri | 1 | 762.98 |
| Total | South | Mississippi | 1 | 587.76 |
| Total | West | Montana | 1 | 480.73 |
| Total | South | North Carolina | 1 | 587.76 |
| Total | Midwest | North Dakota | 1 | 762.98 |
| Total | Midwest | Nebraska | 1 | 762.98 |
| Total | Northeast | New Hampshire | 1 | 700.68 |
| Total | Northeast | New Jersey | 1 | 700.68 |
| Total | West | New Mexico | 1 | 480.73 |
| Total | West | Nevada | 1 | 480.73 |
| Total | Northeast | New York | 1 | 700.68 |
| Total | Midwest | Ohio | 1 | 762.98 |
| Total | South | Oklahoma | 1 | 587.76 |
| Total | West | Oregon | 1 | 480.73 |
| Total | Northeast | Pennsylvania | 1 | 700.68 |
| Total | Northeast | Rhode Island | 1 | 700.68 |
| Total | South | South Carolina | 1 | 587.76 |
| Total | Midwest | South Dakota | 1 | 762.98 |
| Total | South | Tennessee | 1 | 587.76 |
| Total | South | Texas | 1 | 587.76 |
| Total | West | Utah | 1 | 480.73 |
| Total | South | Virginia | 1 | 587.76 |
| Total | Northeast | Vermont | 1 | 700.68 |
| Total | West | Washington | 1 | 480.73 |
| Total | Midwest | Wisconsin | 1 | 762.98 |
| Total | South | West Virginia | 1 | 587.76 |
| Total | West | Wyoming | 1 | 480.73 |
| Incidence | West | Alaska | 1 | 14.15 |
| Incidence | South | Alabama | 1 | 14.84 |
| Incidence | South | Arkansas | 1 | 14.84 |
| Incidence | West | Arizona | 1 | 14.15 |
| Incidence | West | California | 1 | 14.15 |
| Incidence | West | Colorado | 1 | 14.15 |
| Incidence | Northeast | Connecticut | 1 | 18.08 |
| Incidence | South | District of Columbia | 1 | 14.84 |
| Incidence | South | Delaware | 1 | 14.84 |
| Incidence | South | Florida | 1 | 14.84 |
| Incidence | South | Georgia | 1 | 14.84 |
| Incidence | West | Hawaii | 1 | 14.15 |
| Incidence | Midwest | Iowa | 1 | 19.64 |
| Incidence | West | Idaho | 1 | 14.15 |
| Incidence | Midwest | Illinois | 1 | 19.64 |
| Incidence | Midwest | Indiana | 1 | 19.64 |
| Incidence | Midwest | Kansas | 1 | 19.64 |
| Incidence | South | Kentucky | 1 | 14.84 |
| Incidence | South | Louisiana | 1 | 14.84 |
| Incidence | Northeast | Massachusetts | 1 | 18.08 |
| Incidence | South | Maryland | 1 | 14.84 |
| Incidence | Northeast | Maine | 1 | 18.08 |
| Incidence | Midwest | Michigan | 1 | 19.64 |
| Incidence | Midwest | Minnesota | 1 | 19.64 |
| Incidence | Midwest | Missouri | 1 | 19.64 |
| Incidence | South | Mississippi | 1 | 14.84 |
| Incidence | West | Montana | 1 | 14.15 |
| Incidence | South | North Carolina | 1 | 14.84 |
| Incidence | Midwest | North Dakota | 1 | 19.64 |
| Incidence | Midwest | Nebraska | 1 | 19.64 |
| Incidence | Northeast | New Hampshire | 1 | 18.08 |
| Incidence | Northeast | New Jersey | 1 | 18.08 |
| Incidence | West | New Mexico | 1 | 14.15 |
| Incidence | West | Nevada | 1 | 14.15 |
| Incidence | Northeast | New York | 1 | 18.08 |
| Incidence | Midwest | Ohio | 1 | 19.64 |
| Incidence | South | Oklahoma | 1 | 14.84 |
| Incidence | West | Oregon | 1 | 14.15 |
| Incidence | Northeast | Pennsylvania | 1 | 18.08 |
| Incidence | Northeast | Rhode Island | 1 | 18.08 |
| Incidence | South | South Carolina | 1 | 14.84 |
| Incidence | Midwest | South Dakota | 1 | 19.64 |
| Incidence | South | Tennessee | 1 | 14.84 |
| Incidence | South | Texas | 1 | 14.84 |
| Incidence | West | Utah | 1 | 14.15 |
| Incidence | South | Virginia | 1 | 14.84 |
| Incidence | Northeast | Vermont | 1 | 18.08 |
| Incidence | West | Washington | 1 | 14.15 |
| Incidence | Midwest | Wisconsin | 1 | 19.64 |
| Incidence | South | West Virginia | 1 | 14.84 |
| Incidence | West | Wyoming | 1 | 14.15 |
| Total | West | Alaska | 5 | 486.70 |
| Total | South | Alabama | 5 | 578.11 |
| Total | South | Arkansas | 5 | 578.11 |
| Total | West | Arizona | 5 | 486.70 |
| Total | West | California | 5 | 486.70 |
| Total | West | Colorado | 5 | 486.70 |
| Total | Northeast | Connecticut | 5 | 714.99 |
| Total | South | District of Columbia | 5 | 578.11 |
| Total | South | Delaware | 5 | 578.11 |
| Total | South | Florida | 5 | 578.11 |
| Total | South | Georgia | 5 | 578.11 |
| Total | West | Hawaii | 5 | 486.70 |
| Total | Midwest | Iowa | 5 | 785.51 |
| Total | West | Idaho | 5 | 486.70 |
| Total | Midwest | Illinois | 5 | 785.51 |
| Total | Midwest | Indiana | 5 | 785.51 |
| Total | Midwest | Kansas | 5 | 785.51 |
| Total | South | Kentucky | 5 | 578.11 |
| Total | South | Louisiana | 5 | 578.11 |
| Total | Northeast | Massachusetts | 5 | 714.99 |
| Total | South | Maryland | 5 | 578.11 |
| Total | Northeast | Maine | 5 | 714.99 |
| Total | Midwest | Michigan | 5 | 785.51 |
| Total | Midwest | Minnesota | 5 | 785.51 |
| Total | Midwest | Missouri | 5 | 785.51 |
| Total | South | Mississippi | 5 | 578.11 |
| Total | West | Montana | 5 | 486.70 |
| Total | South | North Carolina | 5 | 578.11 |
| Total | Midwest | North Dakota | 5 | 785.51 |
| Total | Midwest | Nebraska | 5 | 785.51 |
| Total | Northeast | New Hampshire | 5 | 714.99 |
| Total | Northeast | New Jersey | 5 | 714.99 |
| Total | West | New Mexico | 5 | 486.70 |
| Total | West | Nevada | 5 | 486.70 |
| Total | Northeast | New York | 5 | 714.99 |
| Total | Midwest | Ohio | 5 | 785.51 |
| Total | South | Oklahoma | 5 | 578.11 |
| Total | West | Oregon | 5 | 486.70 |
| Total | Northeast | Pennsylvania | 5 | 714.99 |
| Total | Northeast | Rhode Island | 5 | 714.99 |
| Total | South | South Carolina | 5 | 578.11 |
| Total | Midwest | South Dakota | 5 | 785.51 |
| Total | South | Tennessee | 5 | 578.11 |
| Total | South | Texas | 5 | 578.11 |
| Total | West | Utah | 5 | 486.70 |
| Total | South | Virginia | 5 | 578.11 |
| Total | Northeast | Vermont | 5 | 714.99 |
| Total | West | Washington | 5 | 486.70 |
| Total | Midwest | Wisconsin | 5 | 785.51 |
| Total | South | West Virginia | 5 | 578.11 |
| Total | West | Wyoming | 5 | 486.70 |
| Incidence | West | Alaska | 5 | 16.35 |
| Incidence | South | Alabama | 5 | 16.46 |
| Incidence | South | Arkansas | 5 | 16.46 |
| Incidence | West | Arizona | 5 | 16.35 |
| Incidence | West | California | 5 | 16.35 |
| Incidence | West | Colorado | 5 | 16.35 |
| Incidence | Northeast | Connecticut | 5 | 20.92 |
| Incidence | South | District of Columbia | 5 | 16.46 |
| Incidence | South | Delaware | 5 | 16.46 |
| Incidence | South | Florida | 5 | 16.46 |
| Incidence | South | Georgia | 5 | 16.46 |
| Incidence | West | Hawaii | 5 | 16.35 |
| Incidence | Midwest | Iowa | 5 | 22.32 |
| Incidence | West | Idaho | 5 | 16.35 |
| Incidence | Midwest | Illinois | 5 | 22.32 |
| Incidence | Midwest | Indiana | 5 | 22.32 |
| Incidence | Midwest | Kansas | 5 | 22.32 |
| Incidence | South | Kentucky | 5 | 16.46 |
| Incidence | South | Louisiana | 5 | 16.46 |
| Incidence | Northeast | Massachusetts | 5 | 20.92 |
| Incidence | South | Maryland | 5 | 16.46 |
| Incidence | Northeast | Maine | 5 | 20.92 |
| Incidence | Midwest | Michigan | 5 | 22.32 |
| Incidence | Midwest | Minnesota | 5 | 22.32 |
| Incidence | Midwest | Missouri | 5 | 22.32 |
| Incidence | South | Mississippi | 5 | 16.46 |
| Incidence | West | Montana | 5 | 16.35 |
| Incidence | South | North Carolina | 5 | 16.46 |
| Incidence | Midwest | North Dakota | 5 | 22.32 |
| Incidence | Midwest | Nebraska | 5 | 22.32 |
| Incidence | Northeast | New Hampshire | 5 | 20.92 |
| Incidence | Northeast | New Jersey | 5 | 20.92 |
| Incidence | West | New Mexico | 5 | 16.35 |
| Incidence | West | Nevada | 5 | 16.35 |
| Incidence | Northeast | New York | 5 | 20.92 |
| Incidence | Midwest | Ohio | 5 | 22.32 |
| Incidence | South | Oklahoma | 5 | 16.46 |
| Incidence | West | Oregon | 5 | 16.35 |
| Incidence | Northeast | Pennsylvania | 5 | 20.92 |
| Incidence | Northeast | Rhode Island | 5 | 20.92 |
| Incidence | South | South Carolina | 5 | 16.46 |
| Incidence | Midwest | South Dakota | 5 | 22.32 |
| Incidence | South | Tennessee | 5 | 16.46 |
| Incidence | South | Texas | 5 | 16.46 |
| Incidence | West | Utah | 5 | 16.35 |
| Incidence | South | Virginia | 5 | 16.46 |
| Incidence | Northeast | Vermont | 5 | 20.92 |
| Incidence | West | Washington | 5 | 16.35 |
| Incidence | Midwest | Wisconsin | 5 | 22.32 |
| Incidence | South | West Virginia | 5 | 16.46 |
| Incidence | West | Wyoming | 5 | 16.35 |
| Total | West | Alaska | 10 | 504.87 |
| Total | South | Alabama | 10 | 575.43 |
| Total | South | Arkansas | 10 | 575.43 |
| Total | West | Arizona | 10 | 504.87 |
| Total | West | California | 10 | 504.87 |
| Total | West | Colorado | 10 | 504.87 |
| Total | Northeast | Connecticut | 10 | 751.45 |
| Total | South | District of Columbia | 10 | 575.43 |
| Total | South | Delaware | 10 | 575.43 |
| Total | South | Florida | 10 | 575.43 |
| Total | South | Georgia | 10 | 575.43 |
| Total | West | Hawaii | 10 | 504.87 |
| Total | Midwest | Iowa | 10 | 826.38 |
| Total | West | Idaho | 10 | 504.87 |
| Total | Midwest | Illinois | 10 | 826.38 |
| Total | Midwest | Indiana | 10 | 826.38 |
| Total | Midwest | Kansas | 10 | 826.38 |
| Total | South | Kentucky | 10 | 575.43 |
| Total | South | Louisiana | 10 | 575.43 |
| Total | Northeast | Massachusetts | 10 | 751.45 |
| Total | South | Maryland | 10 | 575.43 |
| Total | Northeast | Maine | 10 | 751.45 |
| Total | Midwest | Michigan | 10 | 826.38 |
| Total | Midwest | Minnesota | 10 | 826.38 |
| Total | Midwest | Missouri | 10 | 826.38 |
| Total | South | Mississippi | 10 | 575.43 |
| Total | West | Montana | 10 | 504.87 |
| Total | South | North Carolina | 10 | 575.43 |
| Total | Midwest | North Dakota | 10 | 826.38 |
| Total | Midwest | Nebraska | 10 | 826.38 |
| Total | Northeast | New Hampshire | 10 | 751.45 |
| Total | Northeast | New Jersey | 10 | 751.45 |
| Total | West | New Mexico | 10 | 504.87 |
| Total | West | Nevada | 10 | 504.87 |
| Total | Northeast | New York | 10 | 751.45 |
| Total | Midwest | Ohio | 10 | 826.38 |
| Total | South | Oklahoma | 10 | 575.43 |
| Total | West | Oregon | 10 | 504.87 |
| Total | Northeast | Pennsylvania | 10 | 751.45 |
| Total | Northeast | Rhode Island | 10 | 751.45 |
| Total | South | South Carolina | 10 | 575.43 |
| Total | Midwest | South Dakota | 10 | 826.38 |
| Total | South | Tennessee | 10 | 575.43 |
| Total | South | Texas | 10 | 575.43 |
| Total | West | Utah | 10 | 504.87 |
| Total | South | Virginia | 10 | 575.43 |
| Total | Northeast | Vermont | 10 | 751.45 |
| Total | West | Washington | 10 | 504.87 |
| Total | Midwest | Wisconsin | 10 | 826.38 |
| Total | South | West Virginia | 10 | 575.43 |
| Total | West | Wyoming | 10 | 504.87 |
| Incidence | West | Alaska | 10 | 19.58 |
| Incidence | South | Alabama | 10 | 19.32 |
| Incidence | South | Arkansas | 10 | 19.32 |
| Incidence | West | Arizona | 10 | 19.58 |
| Incidence | West | California | 10 | 19.58 |
| Incidence | West | Colorado | 10 | 19.58 |
| Incidence | Northeast | Connecticut | 10 | 26.94 |
| Incidence | South | District of Columbia | 10 | 19.32 |
| Incidence | South | Delaware | 10 | 19.32 |
| Incidence | South | Florida | 10 | 19.32 |
| Incidence | South | Georgia | 10 | 19.32 |
| Incidence | West | Hawaii | 10 | 19.58 |
| Incidence | Midwest | Iowa | 10 | 27.27 |
| Incidence | West | Idaho | 10 | 19.58 |
| Incidence | Midwest | Illinois | 10 | 27.27 |
| Incidence | Midwest | Indiana | 10 | 27.27 |
| Incidence | Midwest | Kansas | 10 | 27.27 |
| Incidence | South | Kentucky | 10 | 19.32 |
| Incidence | South | Louisiana | 10 | 19.32 |
| Incidence | Northeast | Massachusetts | 10 | 26.94 |
| Incidence | South | Maryland | 10 | 19.32 |
| Incidence | Northeast | Maine | 10 | 26.94 |
| Incidence | Midwest | Michigan | 10 | 27.27 |
| Incidence | Midwest | Minnesota | 10 | 27.27 |
| Incidence | Midwest | Missouri | 10 | 27.27 |
| Incidence | South | Mississippi | 10 | 19.32 |
| Incidence | West | Montana | 10 | 19.58 |
| Incidence | South | North Carolina | 10 | 19.32 |
| Incidence | Midwest | North Dakota | 10 | 27.27 |
| Incidence | Midwest | Nebraska | 10 | 27.27 |
| Incidence | Northeast | New Hampshire | 10 | 26.94 |
| Incidence | Northeast | New Jersey | 10 | 26.94 |
| Incidence | West | New Mexico | 10 | 19.58 |
| Incidence | West | Nevada | 10 | 19.58 |
| Incidence | Northeast | New York | 10 | 26.94 |
| Incidence | Midwest | Ohio | 10 | 27.27 |
| Incidence | South | Oklahoma | 10 | 19.32 |
| Incidence | West | Oregon | 10 | 19.58 |
| Incidence | Northeast | Pennsylvania | 10 | 26.94 |
| Incidence | Northeast | Rhode Island | 10 | 26.94 |
| Incidence | South | South Carolina | 10 | 19.32 |
| Incidence | Midwest | South Dakota | 10 | 27.27 |
| Incidence | South | Tennessee | 10 | 19.32 |
| Incidence | South | Texas | 10 | 19.32 |
| Incidence | West | Utah | 10 | 19.58 |
| Incidence | South | Virginia | 10 | 19.32 |
| Incidence | Northeast | Vermont | 10 | 26.94 |
| Incidence | West | Washington | 10 | 19.58 |
| Incidence | Midwest | Wisconsin | 10 | 27.27 |
| Incidence | South | West Virginia | 10 | 19.32 |
| Incidence | West | Wyoming | 10 | 19.58 |

EXHIBIT 3 (figure data)

Data for Distribution of total T1DM and incident T1DM populations by age band in model years 1, 5, and 10

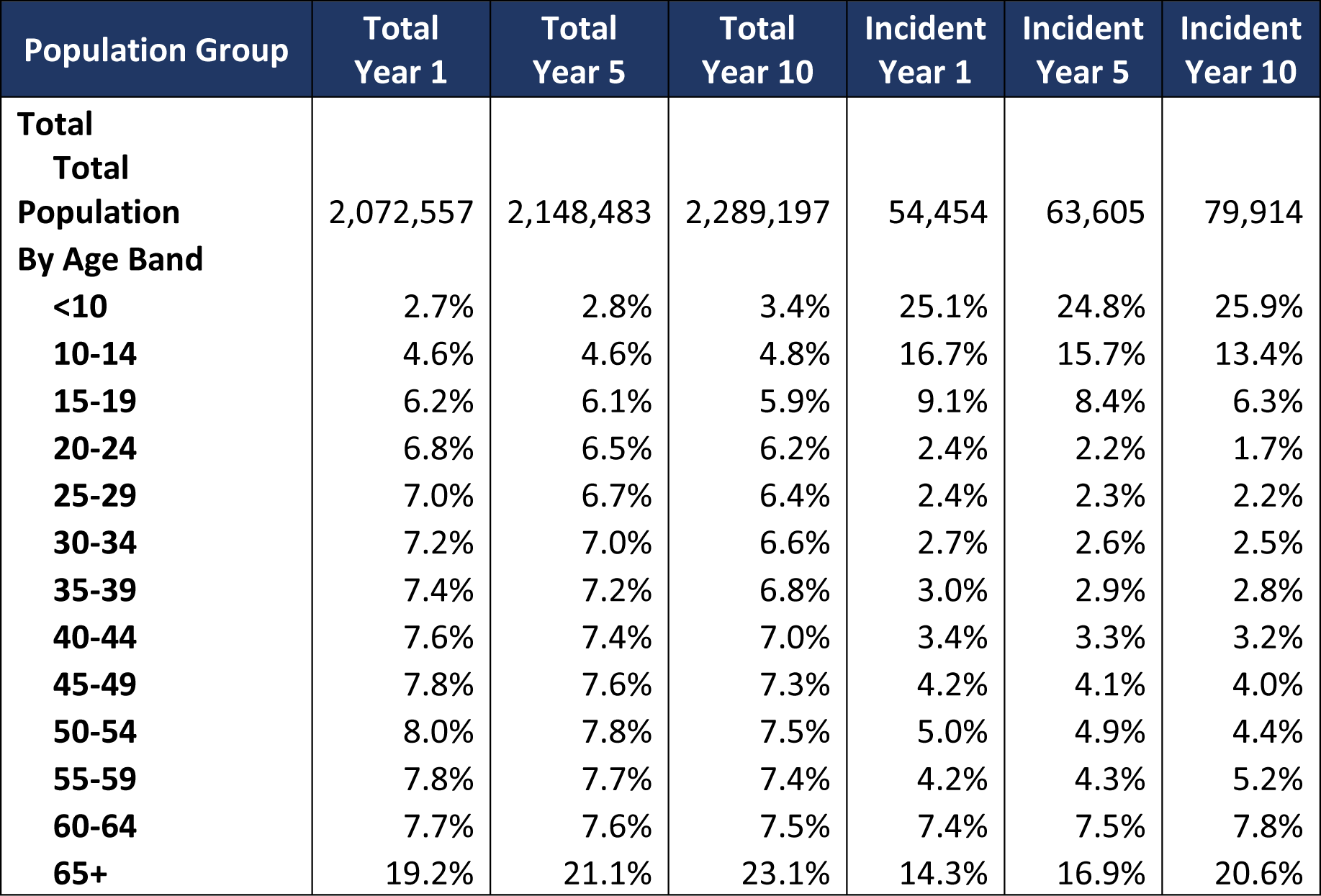

| Population Group | Total<br>Year 1 | Total<br>Year 5 | Total<br>Year 10 | Incident<br>Year 1 | Incident<br>Year 5 | Incident<br>Year 10 |
| --- | --- | --- | --- | --- | --- | --- |
| **Total** |  |  |  |  |  |  |
| **Total**<br>**Population** | 2,072,557 | 2,148,483 | 2,289,197 | 54,454 | 63,605 | 79,914 |
| **By Age Band** |  |  |  |  |  |  |
| **<10** | 2.7% | 2.8% | 3.4% | 25.1% | 24.8% | 25.9% |
| **10-14** | 4.6% | 4.6% | 4.8% | 16.7% | 15.7% | 13.4% |
| **15-19** | 6.2% | 6.1% | 5.9% | 9.1% | 8.4% | 6.3% |
| **20-24** | 6.8% | 6.5% | 6.2% | 2.4% | 2.2% | 1.7% |
| **25-29** | 7.0% | 6.7% | 6.4% | 2.4% | 2.3% | 2.2% |
| **30-34** | 7.2% | 7.0% | 6.6% | 2.7% | 2.6% | 2.5% |
| **35-39** | 7.4% | 7.2% | 6.8% | 3.0% | 2.9% | 2.8% |
| **40-44** | 7.6% | 7.4% | 7.0% | 3.4% | 3.3% | 3.2% |
| **45-49** | 7.8% | 7.6% | 7.3% | 4.2% | 4.1% | 4.0% |
| **50-54** | 8.0% | 7.8% | 7.5% | 5.0% | 4.9% | 4.4% |
| **55-59** | 7.8% | 7.7% | 7.4% | 4.2% | 4.3% | 5.2% |
| **60-64** | 7.7% | 7.6% | 7.5% | 7.4% | 7.5% | 7.8% |
| **65+** | 19.2% | 21.1% | 23.1% | 14.3% | 16.9% | 20.6% |

