## Supplemental Files for "We Are on the Verge of Breakthrough Cures for Type 1 Diabetes, but Who Are the 2 Million Americans Who Have It?"

**APPENDIX**

**Exhibit A1. Population Sizes of Each Market in Data and in the Nation**

| Market | Population Size (in Data) | Population Size in Millions | Source |
|---|---|---|---|
| Commercial | 10,769,405 | 11 | Milliman Consolidated Health Cost Sources Database |
| Medicaid | 4,040,095 | 4 | Milliman Consolidated Health Cost Sources Database |
| Medicare Fee-for-Service | 20,607,550 | 21 | CMS 100% Research Identifiable Medicare Data |
| Medicare Advantage | 20,776,377 | 21 | CMS 100% Research Identifiable Medicare Encounter Data |

| Market | Population Size (in Literature) | Population Size in Millions | Source |
|---|---|---|---|
| Commercial | 172,700,000 | 173 | CDC National Center for Health Statistics |
| Medicaid | 64,896,849 | 65 | CMS Medicaid and CHIP Scorecard |
| Medicare Fee-for-Service | 38,577,012 | 39 | CMS Chronic Condition Warehouse |
| Medicare Advantage | 22,200,000 | 22 | Kaiser Family Foundation |
| Veterans Affairs | 4,236,144 | 4 | US Department of Veterans Affairs |
| Uninsured | 32,800,000 | 33 | CDC National Center for Health Statistics |
| Other Medicare Health Plan Enrollment | 3,622,988 | 4 | CMS Chronic Condition Warehouse |

**Exhibit A2. HCPCS Codes for Insulin, Insulin Pumps, and Insulin-Related DME**

| Code Type | Code | Description | Insulin Type | Include/Exclude from T1DM |
|---|---|---|---|---|
| HCPCS | A4225 | Supplies for external insulin infusion pump, syringe type cartridge, sterile, each | Not insulin | Include - Insulin Pump-Related Supplies |
| HCPCS | A4226 | Supplies for maintenance of insulin infusion pump with dosage rate adjustment using therapeutic contir | Not insulin | Include - Insulin Pump-Related Supplies |
| HCPCS | A4230 | Infusion set for external insulin pump, non needle cannula type | Not insulin | Include - Insulin Pump-Related Supplies |
| HCPCS | A4231 | Infusion set for external insulin pump, needle type | Not insulin | Include - Insulin Pump-Related Supplies |
| HCPCS | A4232 | Syringe with needle for external insulin pump, sterile, 3 cc | Not insulin | Include - Insulin Pump-Related Supplies |
| HCPCS | A9274 | External ambulatory insulin delivery system, disposable, each, includes all supplies and accessories | Short | Include - Disposble Insulin Pump (contains short acting insulin) |
| HCPCS | E0784 | External ambulatory infusion pump, insulin | Not insulin | Include - Insulin Pump |
| HCPCS | E0787 | External ambulatory infusion pump, insulin, dosage rate adjustment using therapeutic continuous gluco | Not insulin | Include - Insulin Pump |
| HCPCS | J1815 | Injection, insulin, per 5 units | Short | Include - Short acting insulin for T1DM ID. May be used in an acute episode for patients without diabetes. |
| HCPCS | J1817 | Insulin for administration through dme (i.e., insulin pump) per 50 units | Short | Include - Short acting insulin for T1DM ID |
| HCPCS | K0601 | Replacement battery for external infusion pump owned by patient, silver oxide, 1.5 volt, each | Not insulin | Include - Insulin Pump-Related Supplies |
| HCPCS | K0602 | Replacement battery for external infusion pump owned by patient, silver oxide, 3 volt, each | Not insulin | Include - Insulin Pump-Related Supplies |
| HCPCS | K0603 | Replacement battery for external infusion pump owned by patient, alkaline, 1.5 volt, each | Not insulin | Include - Insulin Pump-Related Supplies |
| HCPCS | K0604 | Replacement battery for external infusion pump owned by patient, lithium, 3.6 volt, each | Not insulin | Include - Insulin Pump-Related Supplies |
| HCPCS | K0605 | Replacement battery for external infusion pump owned by patient, lithium, 4.5 volt, each | Not insulin | Include - Insulin Pump-Related Supplies |
| HCPCS | S1034 | Artificial pancreas device system (e.g., low glucose suspend (lgs) feature) including continuous glucose monitor, blood glucose device, insulin pump and computer algorithm that communicates with all of the devices | Not insulin | Include - Insulin Pump |
| HCPCS | S1035 | Sensor; invasive (e.g., subcutaneous), disposable, for use with artificial pancreas device system | Not insulin | Include - Insulin Pump-Related Supplies |
| HCPCS | S1036 | Transmitter; External, For Use With Artificial Pancreas Device System | Not insulin | Include - Insulin Pump-Related Supplies |
| HCPCS | S1037 | Receiver (Monitor); External, For Use With Artificial Pancreas Device System | Not insulin | Include - Insulin Pump-Related Supplies |
| HCPCS | S5550 | Insulin, rapid onset, 5 units | Short | Include - Short acting insulin for T1DM ID. May be used in an acute episode for patients without diabetes. |
| HCPCS | S5551 | Insulin, most rapid onset (lispro or aspart); 5 units | Short | Include - Short acting insulin for T1DM ID. May be used in an acute episode for patients without diabetes. |
| HCPCS | S5552 | Insulin, intermediate acting (nph or lente); 5 units | Intermediate | Exclude from T1DM ID - Intermediate acting insulin. May be used in an acute episode for patients without diabetes. |
| HCPCS | S5553 | Insulin, long acting; 5 units | Long | Exclude from T1DM ID - Long acting insulin. May be used in an acute episode for patients without diabetes. |
| HCPCS | S5560 | Insulin delivery device, reusable pen; 1.5 ml size | Not insulin | Include - Insulin-Related Supplies |
| HCPCS | S5561 | Insulin delivery device, reusable pen; 3 ml size | Not insulin | Include - Insulin-Related Supplies |
| HCPCS | S5565 | Insulin cartridge for use in insulin delivery device other than pump; 150 units | Short | Include - Short acting insulin for T1DM ID |
| HCPCS | S5566 | Insulin cartridge for use in insulin delivery device other than pump; 300 units | Short | Include - Short acting insulin for T1DM ID |
| HCPCS | S5570 | Insulin delivery device, disposable pen (including insulin); 1.5 ml size | Short | Include - Short acting insulin for T1DM ID |
| HCPCS | S5571 | Insulin delivery device, disposable pen (including insulin); 3 ml size | Short | Include - Short acting insulin for T1DM ID |
| HCPCS | S8490 | Insulin syringes (100 syringes, any size) | Not insulin | Include - Insulin-Related Supplies |

**Exhibit A3. NDC Codes for Insulin, Insulin Pumps, and Insulin-Related DME**

NDCs were generated for each of the description categories and insulin types listed below. The full list of NDCs is available from the authors.

| Code Type | Description | Insulin Type | Include/Exclude from T1DM |
|---|---|---|---|
| NDC | Insulin Aspart Inj 100 Unit/ML | Short | Include - Short-acting insulin for T1DM ID |
| NDC | Insulin Aspart Soln Pen-injector 100 Unit/ML | Short | Include - Short-acting insulin for T1DM ID |
| NDC | Insulin Aspart Soln Cartridge 100 Unit/ML | Short | Include - Short-acting insulin for T1DM ID |
| NDC | Insulin Glargine Inj 100 Unit/ML | Long | Exclude from T1DM ID - Long acting insulin |
| NDC | Insulin Glargine Soln Pen-Injector 100 Unit/ML | Long | Exclude from T1DM ID - Long acting insulin |
| NDC | Insulin Glargine Soln Pen-Injector 300 Unit/ML | Long | Exclude from T1DM ID - Long acting insulin |
| NDC | Insulin Glulisine Inj 100 Unit/ML | Short | Include - Short-acting insulin for T1DM ID |
| NDC | Insulin Glulisine Soln Pen-Injector Inj 100 Unit/ML | Short | Include - Short-acting insulin for T1DM ID |
| NDC | Insulin Lispro Inj 100 Unit/ML | Short | Include - Short-acting insulin for T1DM ID |
| NDC | Insulin Lispro Soln Pen-injector 100 Unit/ML | Short | Include - Short-acting insulin for T1DM ID |
| NDC | Insulin Lispro Soln Pen-injector 200 Unit/ML | Short | Include - Short-acting insulin for T1DM ID |
| NDC | Insulin Lispro Soln Cartridge 100 Unit/ML | Short | Include - Short-acting insulin for T1DM ID |
| NDC | Insulin Detemir Inj 100 Unit/ML | Long | Exclude from T1DM ID - Long acting insulin |
| NDC | Insulin Detemir Soln Pen-injector 100 Unit/ML | Long | Exclude from T1DM ID - Long acting insulin |
| NDC | Insulin Degludec Inj 100 Unit/ML | Long | Exclude from T1DM ID - Long acting insulin |
| NDC | Insulin Degludec Soln Pen-Injector 100 Unit/ML | Long | Exclude from T1DM ID - Long acting insulin |
| NDC | Insulin Degludec Soln Pen-Injector 200 Unit/ML | Long | Exclude from T1DM ID - Long acting insulin |
| NDC | Insulin Regular (Human) Inj 100 Unit/ML | Short | Include - Short-acting insulin for T1DM ID |
| NDC | Insulin Regular (Human) Inj 500 Unit/ML | Short | Include - Short-acting insulin for T1DM ID |
| NDC | Insulin Regular (Human) Inhalation Powder 4 Unit/Cartridge | Short | Include - Short-acting insulin for T1DM ID |
| NDC | Insulin Regular (Human) Inhalation Powder 8 Unit/Cartridge | Short | Include - Short-acting insulin for T1DM ID |
| NDC | Insulin Regular (Human) Inhalation Powder 12 Unit/Cartridge | Short | Include - Short-acting insulin for T1DM ID |
| NDC | Insulin Regular (Human) Inhal Powd 4 (30) & 8 (60) Unit/Cart | Short | Include - Short-acting insulin for T1DM ID |
| NDC | Insulin Regular (Human) Inhal Powd 4 (60) & 8 (30) Unit/Cart | Short | Include - Short-acting insulin for T1DM ID |
| NDC | Insulin Regular (Human) Inhal Powd 4 (90) & 8 (90) Unit/Cart | Short | Include - Short-acting insulin for T1DM ID |
| NDC | Insulin Regular (Human) Inh Powd 8 (60) & 12 (30) Unit/Cart | Short | Include - Short-acting insulin for T1DM ID |
| NDC | Insulin Regular (Human) Inh Powd 8 (90) & 12 (90) Unit/Cart | Short | Include - Short-acting insulin for T1DM ID |
| NDC | Insulin Regular (Human) Inh Powd 4 & 8 & 12 Unit/Cart (60) | Short | Include - Short-acting insulin for T1DM ID |
| NDC | Insulin Regular (Human) Soln Pen-Injector 500 Unit/ML | Short | Include - Short-acting insulin for T1DM ID |
| NDC | Insulin Isophane (Human) Inj 100 Unit/ML | Short | Include - Short-acting insulin for T1DM ID |
| NDC | Insulin NPH (Human) (Isophane) Inj 100 Unit/ML | Intermediate | Include - Intermediate-acting insulin for T1DM ID |
| NDC | Insulin NPH (Human) (Isophane) Susp Pen-injector 100 Unit/ML | Intermediate | Include - Intermediate-acting insulin for T1DM ID |
| NDC | Insulin Aspart Prot & Aspart (Human) Inj 100 Unit/ML (70-30) | Short, Intermediate | Include - Short-acting insulin for T1DM ID |
| NDC | Insulin Aspart Prot & Aspart Sus Pen-inj 100 Unit/ML (70-30) | Short, Intermediate | Include - Short-acting insulin for T1DM ID |
| NDC | Insulin Lispro Prot & Lispro Inj 100 Unit/ML (75-25) | Short, Intermediate | Include - Short-acting insulin for T1DM ID |
| NDC | Insulin Lispro Protamine & Lispro Inj 100 Unit/ML (50-50) | Short, Intermediate | Include - Short-acting insulin for T1DM ID |
| NDC | Insulin Lispro Prot & Lispro Sus Pen-inj 100 Unit/ML (75-25) | Short, Intermediate | Include - Short-acting insulin for T1DM ID |
| NDC | Insulin Lispro Prot & Lispro Sus Pen-inj 100 Unit/ML (50-50) | Short, Intermediate | Include - Short-acting insulin for T1DM ID |
| NDC | Insulin Isophane & Regular (Human) Inj 100 Unit/ML (70-30) | Short, Intermediate | Include - Short-acting insulin for T1DM ID |
| NDC | Insulin NPH Isophane & Regular Human Inj 100 Unit/ML (70-30) | Short, Intermediate | Include - Short-acting insulin for T1DM ID |
| NDC | Insulin NPH & Regular Susp Pen-Inj 100 Unit/ML (70-30) | Short, Intermediate | Include - Short-acting insulin for T1DM ID |
| NDC | Insulin Degludec-Liraglutide Sol Pen-Inj 100-3.6 Unit-MG/ML | Long, GLP1 | Exclude from T1DM ID - Long acting insulin |
| NDC | Insulin Glargine-Lixisenatide Sol Pen-Inj 100-33 Unit-MCG/ML | Long, GLP1 | Exclude from T1DM ID - Long acting insulin |
| NDC | Insulin Syringe (Disp) U-100 0.3 ML | Not insulin | Include - Insulin-Related Supplies |
| NDC | Insulin Syringe (Disp) U-100 1/2 ML | Not insulin | Include - Insulin-Related Supplies |
| NDC | Insulin Syringe (Disp) U-100 1 ML | Not insulin | Include - Insulin-Related Supplies |
| NDC | Insulin Syringe/Needle U-40 1 ML 25 x 5/8" | Not insulin | Include - Insulin-Related Supplies |
| NDC | Insulin Syringe/Needle U-100 0.3 ML 29 G | Not insulin | Include - Insulin-Related Supplies |
| NDC | Insulin Syringe/Needle U-100 0.3 ML 30 G | Not insulin | Include - Insulin-Related Supplies |
| NDC | Insulin Syringe/Needle U-100 0.3 ML 28 x 1/2" | Not insulin | Include - Insulin-Related Supplies |
| NDC | Insulin Syringe/Needle U-100 0.3 ML 29 x 1/2" | Not insulin | Include - Insulin-Related Supplies |
| NDC | Insulin Syringe/Needle U-100 0.3 ML 30 x 3/8" | Not insulin | Include - Insulin-Related Supplies |
| NDC | Insulin Syringe/Needle U-100 0.3 ML 30 x 5/16" | Not insulin | Include - Insulin-Related Supplies |
| NDC | Insulin Syringe/Needle U-100 0.3 ML 30 x 1/2" | Not insulin | Include - Insulin-Related Supplies |
| NDC | Insulin Syringe/Needle U-100 1/2 ML 27 x 1/2" | Not insulin | Include - Insulin-Related Supplies |
| NDC | Insulin Syringe/Needle U-100 1/2 ML 29 G | Not insulin | Include - Insulin-Related Supplies |
| NDC | Insulin Syringe/Needle U-100 1/2 ML 30 G | Not insulin | Include - Insulin-Related Supplies |
| NDC | Insulin Syringe/Needle U-100 1/2 ML 30 x 3/8" | Not insulin | Include - Insulin-Related Supplies |
| NDC | Insulin Syringe/Needle U-100 1/2 ML 31 x 5/16" | Not insulin | Include - Insulin-Related Supplies |
| NDC | Insulin Syringe/Needle U-100 1/2 ML 28 x 5/16" | Not insulin | Include - Insulin-Related Supplies |
| NDC | Insulin Syringe/Needle U-100 1/2 ML 28 x 1/2" | Not insulin | Include - Insulin-Related Supplies |
| NDC | Insulin Syringe/Needle U-100 1/2 ML 29 x 5/16" | Not insulin | Include - Insulin-Related Supplies |
| NDC | Insulin Syringe/Needle U-100 1/2 ML 29 x 1/2" | Not insulin | Include - Insulin-Related Supplies |
| NDC | Insulin Syringe/Needle U-100 1/2 ML 30 x 5/16" | Not insulin | Include - Insulin-Related Supplies |
| NDC | Insulin Syringe/Needle U-100 1/2 ML 30 x 1/2" | Not insulin | Include - Insulin-Related Supplies |
| NDC | Insulin Syringe/Needle U-100 1 ML 25 x 5/8" | Not insulin | Include - Insulin-Related Supplies |
| NDC | Insulin Syringe/Needle U-100 1/2 ML 31 x 3/8" | Not insulin | Include - Insulin-Related Supplies |
| NDC | Insulin Syringe/Needle U-100 1 ML 31 x 3/8" | Not insulin | Include - Insulin-Related Supplies |
| NDC | Insulin Syringe/Needle U-100 0.3 ML 31 x 15/64" | Not insulin | Include - Insulin-Related Supplies |
| NDC | Insulin Syringe/Needle U-100 0.3 ML 31 x 1/4" (6 MM) | Not insulin | Include - Insulin-Related Supplies |
| NDC | Insulin Syringe/Needle U-100 1 ML 25 x 1" | Not insulin | Include - Insulin-Related Supplies |
| NDC | Insulin Syringe/Needle U-100 0.5 ML 31 x 1/4" (6 MM) | Not insulin | Include - Insulin-Related Supplies |
| NDC | Insulin Syringe/Needle U-100 1 ML 31 x 1/4" (6 MM) | Not insulin | Include - Insulin-Related Supplies |
| NDC | Insulin Syringe/Needle U-100 1 ML 30 x 3/16" (5 MM) | Not insulin | Include - Insulin-Related Supplies |
| NDC | Insulin Syringe/Needle U-100 1 ML 26 x 1/2" | Not insulin | Include - Insulin-Related Supplies |
| NDC | Insulin Syringe/Needle U-100 1 ML 27 x 1/2" | Not insulin | Include - Insulin-Related Supplies |
| NDC | Insulin Syringe/Needle U-100 0.5 ML 30 x 3/16" (5 MM) | Not insulin | Include - Insulin-Related Supplies |
| NDC | Insulin Syringe/Needle U-100 0.3 ML 30 x 15/64" | Not insulin | Include - Insulin-Related Supplies |
| NDC | Insulin Syringe/Needle U-100 1 ML 27 x 5/8" | Not insulin | Include - Insulin-Related Supplies |
| NDC | Insulin Syringe/Needle U-100 0.5 ML 30 x 15/64" | Not insulin | Include - Insulin-Related Supplies |
| NDC | Insulin Syringe/Needle U-100 1 ML 30 x 15/64" | Not insulin | Include - Insulin-Related Supplies |

**Exhibit A3. NDC Codes for Insulin, Insulin Pumps, and Insulin-Related DME**

NDCs were generated for each of the description categories and insulin types listed below. The full list of NDCs is available from the authors.

| Code Type | Description | Insulin Type | Include/Exclude from T1DM |
|---|---|---|---|
| NDC | Insulin Aspart Inj 100 Unit/ML | Short | Include - Short-acting insulin for T1DM ID |
| NDC | Insulin Syringe/Needle U-100 1 ML 28 x 5/16" | Not insulin | Include - Insulin-Related Supplies |
| NDC | Insulin Syringe/Needle U-100 1 ML 28 x 1/2" | Not insulin | Include - Insulin-Related Supplies |
| NDC | Insulin Syringe/Needle U-100 1 ML 29 G | Not insulin | Include - Insulin-Related Supplies |
| NDC | Insulin Syringe/Needle U-100 1 ML 29 x 1/2" | Not insulin | Include - Insulin-Related Supplies |
| NDC | Insulin Syringe/Needle U-100 1 ML 29 x 5/16" | Not insulin | Include - Insulin-Related Supplies |
| NDC | Insulin Syringe/Needle U-100 1 ML 30 G | Not insulin | Include - Insulin-Related Supplies |
| NDC | Insulin Syringe/Needle U-100 1 ML 30 x 5/16" | Not insulin | Include - Insulin-Related Supplies |
| NDC | Insulin Syringe/Needle U-100 1 ML 30 x 1/2" | Not insulin | Include - Insulin-Related Supplies |
| NDC | Insulin Syringe/Needle U-100 1 ML 31 x 5/16" | Not insulin | Include - Insulin-Related Supplies |
| NDC | Insulin Syringe/Needle U-100 0.3 ML 31 x 5/16" | Not insulin | Include - Insulin-Related Supplies |
| NDC | Insulin Syringe/Needle U-100 0.3 ML 31 x 3/8" | Not insulin | Include - Insulin-Related Supplies |
| NDC | Insulin Syringe/Needle U-100 2 ML 27.5 x 5/8" | Not insulin | Include - Insulin-Related Supplies |
| NDC | Insulin Syringe/Needle U-100 1/2 ML 31 x 15/64" | Not insulin | Include - Insulin-Related Supplies |
| NDC | Insulin Syringe/Needle U-100 2 ML 29 x 1/2" | Not insulin | Include - Insulin-Related Supplies |
| NDC | Insulin Syringe/Needle U-100 1 ML 30 x 3/8" | Not insulin | Include - Insulin-Related Supplies |
| NDC | Insulin Syringe/Needle U-100 0.3 ML 29 x 1" | Not insulin | Include - Insulin-Related Supplies |
| NDC | Insulin Syringe/Needle U-100 1 ML 31 x 15/64" | Not insulin | Include - Insulin-Related Supplies |
| NDC | Insulin Syringe/Needle U-500 0.5 ML 31G x 6MM (15/64") | Not insulin | Include - Insulin-Related Supplies |
| NDC | Injection Device - Misc | Short | Include - Short-acting insulin for T1DM ID |
| NDC | Injection Device for Insulin | Not insulin | Include - Insulin-Related Supplies |
| NDC | Injection Device for Insulin | Short | Include - Short-acting insulin for T1DM ID |
| NDC | Insulin Pen Needle 29 G X 5 MM (3/16") | Not insulin | Include - Insulin-Related Supplies |
| NDC | Insulin Pen Needle 29 G X 8 MM (5/16") | Not insulin | Include - Insulin-Related Supplies |
| NDC | Insulin Pen Needle 29 G X 10 MM | Not insulin | Include - Insulin-Related Supplies |
| NDC | Insulin Pen Needle 29 G X 12 MM (1/2") | Not insulin | Include - Insulin-Related Supplies |
| NDC | Insulin Pen Needle 29 G X 12.7 MM | Not insulin | Include - Insulin-Related Supplies |
| NDC | Insulin Pen Needle 29 G X 13 MM (1/2") | Not insulin | Include - Insulin-Related Supplies |
| NDC | Insulin Pen Needle 30 G X 5 MM (3/16") | Not insulin | Include - Insulin-Related Supplies |
| NDC | Insulin Pen Needle 30 G X 8 MM (1/3" or 5/16") | Not insulin | Include - Insulin-Related Supplies |
| NDC | Insulin Pen Needle 31 G X 4 MM (1/6") | Not insulin | Include - Insulin-Related Supplies |
| NDC | Insulin Pen Needle 31 G X 5 MM (3/16") | Not insulin | Include - Insulin-Related Supplies |
| NDC | Insulin Pen Needle 31 G X 6 MM (1/4") | Not insulin | Include - Insulin-Related Supplies |
| NDC | Insulin Pen Needle 31 G X 8 MM (1/3" or 5/16") | Not insulin | Include - Insulin-Related Supplies |
| NDC | Insulin Pen Needle 32 G X 4 MM (5/32") | Not insulin | Include - Insulin-Related Supplies |
| NDC | Insulin Pen Needle 32 G X 5 MM (1/5" or 3/16") | Not insulin | Include - Insulin-Related Supplies |
| NDC | Insulin Pen Needle 32 G X 6 MM (1/4") | Not insulin | Include - Insulin-Related Supplies |
| NDC | Insulin Pen Needle 32 G X 8 MM | Not insulin | Include - Insulin-Related Supplies |
| NDC | Insulin Pen Needle 33 G X 4 MM (5/32") | Not insulin | Include - Insulin-Related Supplies |
| NDC | Insulin Pen Needle 33 G X 5 MM (1/5" or 3/16") | Not insulin | Include - Insulin-Related Supplies |
| NDC | Insulin Pen Needle 33 G X 6 MM (1/4") | Not insulin | Include - Insulin-Related Supplies |
| NDC | Insulin Pen Needle 33 G X 8 MM (1/3" or 5/16") | Not insulin | Include - Insulin-Related Supplies |
| NDC | Injection Device Needle-Free for Insulin | Not insulin | Include - Insulin-Related Supplies |
| NDC | Injection Device Needle-Free for Insulin Supplies | Not insulin | Include - Insulin-Related Supplies |
| NDC | Injection Device Needle-Free for Insulin Kit | Not insulin | Include - Insulin-Related Supplies |
| NDC | Insulin Administration Supplies - Misc | Not insulin | Include - Insulin-Related Supplies |
| NDC | Insulin Administration Supplies - Kit | Not insulin | Include - Insulin-Related Supplies |
| NDC | Insulin Infusion Pump - Device | Not insulin | Include - Insulin Pump |
| NDC | Insulin Infusion Pump - Kit | Not insulin | Include - Insulin Pump |
| NDC | Insulin Infusion Pump - Accessories | Not insulin | Include - Insulin Pump-Related Supplies |
| NDC | Insulin Infusion Pump Supplies | Not insulin | Include - Insulin Pump-Related Supplies |
| NDC | Insulin Infusion Disposable Pump Supplies | Short | Include - Disposable Insulin Pump (contains short-acting insulin) |
| NDC | Insulin Infusion Disposable Pump Kit | Short | Include - Disposable Insulin Pump (contains short-acting insulin) |
| NDC | Insulin Aspart (with Niacinamide) Soln Cartridge 100 Unit/ML | Short | Include - Short-acting insulin for T1DM ID |
| NDC | Insulin Glargine Soln Pen-Injector 300 Unit/ML (1 Unit Dial) | Long | Exclude from T1DM ID - Long acting insulin |
| NDC | Insulin Glargine Soln Pen-Injector 300 Unit/ML (2 Unit Dial) | Long | Exclude from T1DM ID - Long acting insulin |
| NDC | Insulin Lispro Soln Pen-injector 100 Unit/ML (0.5 Unit Dial) | Short | Include - Short-acting insulin for T1DM ID |
| NDC | Insulin Lispro Soln Pen-injector 100 Unit/ML (1 Unit Dial) | Short | Include - Short-acting insulin for T1DM ID |
| NDC | Insulin Regular (Human) Soln Pen-Injector 100 Unit/ML | Short | Include - Short-acting insulin for T1DM ID |
| NDC | Insulin Regular (Human) in NaCl 0.9% IV Soln 100 Unit/100ML | Short | Include - Short-acting insulin for T1DM ID |
| NDC | *Injection Device - Kit*** | Short | Include - Insulin patch (incuding short acting insulin so used for T1DM ID) |
| NDC | Injection Device for Insulin | Short | Include - Insulin patch (incuding short acting insulin so used for T1DM ID) |
| NDC | *Injection Device for Insulin - Accessories*** | Short | Include - Insulin patch (incuding short acting insulin so used for T1DM ID) |
| NDC | Insulin Pen Needle 29 G X 5 MM (1/5" or 3/16") | Not insulin | Include - Insulin-Related Supplies |
| NDC | Insulin Pen Needle 29 G X 8 MM (1/3" or 5/16") | Not insulin | Include - Insulin-Related Supplies |
| NDC | Insulin Pen Needle 29 G X 12.7 MM (1/2") | Not insulin | Include - Insulin-Related Supplies |
| NDC | Insulin Pen Needle 30 G X 5 MM (1/5" or 3/16") | Not insulin | Include - Insulin-Related Supplies |
| NDC | Insulin Pen Needle 31 G X 5 MM (1/5" or 3/16") | Not insulin | Include - Insulin-Related Supplies |
| NDC | Insulin Pen Needle 31 G X 6 MM (1/4" or 15/64") | Not insulin | Include - Insulin-Related Supplies |
| NDC | Insulin Pen Needle 32 G X 4 MM (1/6" or 5/32") | Not insulin | Include - Insulin-Related Supplies |
| NDC | Insulin Pen Needle 32 G X 6 MM (1/4" or 15/64") | Not insulin | Include - Insulin-Related Supplies |
| NDC | Insulin Pen Needle 32 G X 8 MM (1/3" or 5/16") | Not insulin | Include - Insulin-Related Supplies |
| NDC | Insulin Pen Needle 33 G X 4 MM (1/6" or 5/32") | Not insulin | Include - Insulin-Related Supplies |
| NDC | Insulin Pen Needle 33 G X 6 MM (1/4" or 15/64") | Not insulin | Include - Insulin-Related Supplies |
| NDC | *Insulin Infusion Pump - Device*** | Not insulin | Include - Insulin Pump |
| NDC | *Insulin Infusion Pump Supplies*** | Not insulin | Include - Insulin Pump-Related Supplies |
| NDC | Insulin Aspart (with Niacinamide) Inj 100 Unit/ML | Short | Include - Short-acting insulin for T1DM ID |
| NDC | Insulin Aspart (with Niacinamide) Sol Pen-inj 100 Unit/ML | Short | Include - Short-acting insulin for T1DM ID |
| NDC | Insulin Lispro-aabc Inj 100 Unit/ML | Short | Include - Short-acting insulin for T1DM ID |
| NDC | Insulin Lispro-aabc Soln Pen-inj 100 Unit/ML (1 Unit Dial) | Short | Include - Short-acting insulin for T1DM ID |
| NDC | Insulin Lispro-aabc Soln Pen-injector 200 Unit/ML | Short | Include - Short-acting insulin for T1DM ID |
| NDC | Insulin Regular (Human) Inhal Powd 30 x 4 Unit & 60 x 8 Unit | Short | Include - Short-acting insulin for T1DM ID |

**Exhibit A3. NDC Codes for Insulin, Insulin Pumps, and Insulin-Related DME**

NDCs were generated for each of the description categories and insulin types listed below. The full list of NDCs is available from the authors.

| Code Type | Description | Insulin Type | Include/Exclude from T1DM |
|---|---|---|---|
| NDC | Insulin Aspart Inj 100 Unit/ML | Short | Include - Short-acting insulin for T1DM ID |
| NDC | Insulin Regular (Human) Inhal Powd 60 x 4 Unit & 30 x 8 Unit | Short | Include - Short-acting insulin for T1DM ID |
| NDC | Insulin Regular (Human) Inhal Powd 90 x 4 Unit & 90 x 8 Unit | Short | Include - Short-acting insulin for T1DM ID |
| NDC | Insulin Regular (Human) Inh Powd 60 x 8 Unit & 30 x 12 Unit | Short | Include - Short-acting insulin for T1DM ID |
| NDC | Insulin Regular (Human) Inh Powd 90 x 8 Unit & 90 x 12 Unit | Short | Include - Short-acting insulin for T1DM ID |
| NDC | Insulin Syringe/Needle U-100 0.5 ML 32 x 5/16" | Not insulin | Include - Insulin-Related Supplies |
| NDC | Insulin Syringe/Needle U-100 1 ML 32 x 5/16" | Not insulin | Include - Insulin-Related Supplies |
| NDC | *Injection Device for Insulin Kit*** | Short | Include - Insulin patch (incuding short acting insulin so used for T1DM ID) |
| NDC | Insulin Pen Needle 31 G X 4 MM (1/6" or 5/32") | Not insulin | Include - Insulin-Related Supplies |
| NDC | Insulin Pen Needle 34 G X 3.5 MM (9/64") | Not insulin | Include - Insulin-Related Supplies |
| NDC | Insulin Infusion Pump Supplies - Infusion Set | Not insulin | Include - Insulin Pump-Related Supplies |
| NDC | Insulin Infusion Pump Supplies - Reservoir | Not insulin | Include - Insulin Pump-Related Supplies |

**Exhibit A4. ICD-10-CM Diagnosis Codes for Type 1 Diabetes**

| Code Type | Code | Description |
|---|---|---|
| ICD-10-CM | E1010 | Type 1 diabetes mellitus with ketoacidosis without coma |
| ICD-10-CM | E1011 | Type 1 diabetes mellitus with ketoacidosis with coma |
| ICD-10-CM | E1021 | Type 1 diabetes mellitus with diabetic nephropathy |
| ICD-10-CM | E1022 | Type 1 diabetes mellitus with diabetic chronic kidney disease |
| ICD-10-CM | E1029 | Type 1 diabetes mellitus with other diabetic kidney complication |
| ICD-10-CM | E10311 | Type 1 diabetes mellitus with unspecified diabetic retinopathy with macular edema |
| ICD-10-CM | E10319 | Type 1 diabetes mellitus with unspecified diabetic retinopathy without macular edema |
| ICD-10-CM | E10321 | Type 1 diabetes mellitus with mild nonproliferative diabetic retinopathy with macular edema |
| ICD-10-CM | E103211 | Type 1 diabetes mellitus with mild nonproliferative diabetic retinopathy with macular edema, right eye |
| ICD-10-CM | E103212 | Type 1 diabetes mellitus with mild nonproliferative diabetic retinopathy with macular edema, left eye |
| ICD-10-CM | E103213 | Type 1 diabetes mellitus with mild nonproliferative diabetic retinopathy with macular edema, bilateral |
| ICD-10-CM | E103219 | Type 1 diabetes mellitus with mild nonproliferative diabetic retinopathy with macular edema, unspecified eye |
| ICD-10-CM | E10329 | Type 1 diabetes mellitus with mild nonproliferative diabetic retinopathy without macular edema |
| ICD-10-CM | E103291 | Type 1 diabetes mellitus with mild nonproliferative diabetic retinopathy without macular edema, right eye |
| ICD-10-CM | E103292 | Type 1 diabetes mellitus with mild nonproliferative diabetic retinopathy without macular edema, left eye |
| ICD-10-CM | E103293 | Type 1 diabetes mellitus with mild nonproliferative diabetic retinopathy without macular edema, bilateral |
| ICD-10-CM | E103299 | Type 1 diabetes mellitus with mild nonproliferative diabetic retinopathy without macular edema, unspecified eye |
| ICD-10-CM | E10331 | Type 1 diabetes mellitus with moderate nonproliferative diabetic retinopathy with macular edema |
| ICD-10-CM | E103311 | Type 1 diabetes mellitus with moderate nonproliferative diabetic retinopathy with macular edema, right eye |
| ICD-10-CM | E103312 | Type 1 diabetes mellitus with moderate nonproliferative diabetic retinopathy with macular edema, left eye |
| ICD-10-CM | E103313 | Type 1 diabetes mellitus with moderate nonproliferative diabetic retinopathy with macular edema, bilateral |
| ICD-10-CM | E103319 | Type 1 diabetes mellitus with moderate nonproliferative diabetic retinopathy with macular edema, unspecified eye |
| ICD-10-CM | E10339 | Type 1 diabetes mellitus with moderate nonproliferative diabetic retinopathy without macular edema |
| ICD-10-CM | E103391 | Type 1 diabetes mellitus with moderate nonproliferative diabetic retinopathy without macular edema, right eye |
| ICD-10-CM | E103392 | Type 1 diabetes mellitus with moderate nonproliferative diabetic retinopathy without macular edema, left eye |
| ICD-10-CM | E103393 | Type 1 diabetes mellitus with moderate nonproliferative diabetic retinopathy without macular edema, bilateral |
| ICD-10-CM | E103399 | Type 1 diabetes mellitus with moderate nonproliferative diabetic retinopathy without macular edema, unspecified eye |
| ICD-10-CM | E10341 | Type 1 diabetes mellitus with severe nonproliferative diabetic retinopathy with macular edema |
| ICD-10-CM | E103411 | Type 1 diabetes mellitus with severe nonproliferative diabetic retinopathy with macular edema, right eye |
| ICD-10-CM | E103412 | Type 1 diabetes mellitus with severe nonproliferative diabetic retinopathy with macular edema, left eye |
| ICD-10-CM | E103413 | Type 1 diabetes mellitus with severe nonproliferative diabetic retinopathy with macular edema, bilateral |
| ICD-10-CM | E103419 | Type 1 diabetes mellitus with severe nonproliferative diabetic retinopathy with macular edema, unspecified eye |
| ICD-10-CM | E10349 | Type 1 diabetes mellitus with severe nonproliferative diabetic retinopathy without macular edema |
| ICD-10-CM | E103491 | Type 1 diabetes mellitus with severe nonproliferative diabetic retinopathy without macular edema, right eye |
| ICD-10-CM | E103492 | Type 1 diabetes mellitus with severe nonproliferative diabetic retinopathy without macular edema, left eye |
| ICD-10-CM | E103493 | Type 1 diabetes mellitus with severe nonproliferative diabetic retinopathy without macular edema, bilateral |
| ICD-10-CM | E103499 | Type 1 diabetes mellitus with severe nonproliferative diabetic retinopathy without macular edema, unspecified eye |
| ICD-10-CM | E10351 | Type 1 diabetes mellitus with proliferative diabetic retinopathy with macular edema |
| ICD-10-CM | E103511 | Type 1 diabetes mellitus with proliferative diabetic retinopathy with macular edema, right eye |
| ICD-10-CM | E103512 | Type 1 diabetes mellitus with proliferative diabetic retinopathy with macular edema, left eye |
| ICD-10-CM | E103513 | Type 1 diabetes mellitus with proliferative diabetic retinopathy with macular edema, bilateral |
| ICD-10-CM | E103519 | Type 1 diabetes mellitus with proliferative diabetic retinopathy with macular edema, unspecified eye |
| ICD-10-CM | E103521 | Type 1 diabetes mellitus with proliferative diabetic retinopathy with traction retinal detachment involving the macula, right eye |
| ICD-10-CM | E103522 | Type 1 diabetes mellitus with proliferative diabetic retinopathy with traction retinal detachment involving the macula, left eye |
| ICD-10-CM | E103523 | Type 1 diabetes mellitus with proliferative diabetic retinopathy with traction retinal detachment involving the macula, bilateral |
| ICD-10-CM | E103529 | Type 1 diabetes mellitus with proliferative diabetic retinopathy with traction retinal detachment involving the macula, unspecified eye |
| ICD-10-CM | E103531 | Type 1 diabetes mellitus with proliferative diabetic retinopathy with traction retinal detachment not involving the macula, right eye |
| ICD-10-CM | E103532 | Type 1 diabetes mellitus with proliferative diabetic retinopathy with traction retinal detachment not involving the macula, left eye |
| ICD-10-CM | E103533 | Type 1 diabetes mellitus with proliferative diabetic retinopathy with traction retinal detachment not involving the macula, bilateral |
| ICD-10-CM | E103539 | Type 1 diabetes mellitus with proliferative diabetic retinopathy with traction retinal detachment not involving the macula, unspecified eye |
| ICD-10-CM | E103541 | Type 1 diabetes mellitus with proliferative diabetic retinopathy with combined traction retinal detachment and rhegmatogenous retinal detachment, right eye |
| ICD-10-CM | E103542 | Type 1 diabetes mellitus with proliferative diabetic retinopathy with combined traction retinal detachment and rhegmatogenous retinal detachment, left eye |
| ICD-10-CM | E103543 | Type 1 diabetes mellitus with proliferative diabetic retinopathy with combined traction retinal detachment and rhegmatogenous retinal detachment, bilateral |
| ICD-10-CM | E103549 | Type 1 diabetes mellitus with proliferative diabetic retinopathy with combined traction retinal detachment and rhegmatogenous retinal detachment, unspecified eye |
| ICD-10-CM | E103551 | Type 1 diabetes mellitus with stable proliferative diabetic retinopathy, right eye |
| ICD-10-CM | E103552 | Type 1 diabetes mellitus with stable proliferative diabetic retinopathy, left eye |
| ICD-10-CM | E103553 | Type 1 diabetes mellitus with stable proliferative diabetic retinopathy, bilateral |
| ICD-10-CM | E103559 | Type 1 diabetes mellitus with stable proliferative diabetic retinopathy, unspecified eye |
| ICD-10-CM | E10359 | Type 1 diabetes mellitus with proliferative diabetic retinopathy without macular edema |
| ICD-10-CM | E103591 | Type 1 diabetes mellitus with proliferative diabetic retinopathy without macular edema, right eye |
| ICD-10-CM | E103592 | Type 1 diabetes mellitus with proliferative diabetic retinopathy without macular edema, left eye |
| ICD-10-CM | E103593 | Type 1 diabetes mellitus with proliferative diabetic retinopathy without macular edema, bilateral |
| ICD-10-CM | E103599 | Type 1 diabetes mellitus with proliferative diabetic retinopathy without macular edema, unspecified eye |
| ICD-10-CM | E1036 | Type 1 diabetes mellitus with diabetic cataract |
| ICD-10-CM | E1037X1 | Type 1 diabetes mellitus with diabetic macular edema, resolved following treatment, right eye |
| ICD-10-CM | E1037X2 | Type 1 diabetes mellitus with diabetic macular edema, resolved following treatment, left eye |
| ICD-10-CM | E1037X3 | Type 1 diabetes mellitus with diabetic macular edema, resolved following treatment, bilateral |
| ICD-10-CM | E1037X9 | Type 1 diabetes mellitus with diabetic macular edema, resolved following treatment, unspecified eye |
| ICD-10-CM | E1039 | Type 1 diabetes mellitus with other diabetic ophthalmic complication |
| ICD-10-CM | E1040 | Type 1 diabetes mellitus with diabetic neuropathy, unspecified |
| ICD-10-CM | E1041 | Type 1 diabetes mellitus with diabetic mononeuropathy |
| ICD-10-CM | E1042 | Type 1 diabetes mellitus with diabetic polyneuropathy |
| ICD-10-CM | E1043 | Type 1 diabetes mellitus with diabetic autonomic (poly)neuropathy |
| ICD-10-CM | E1044 | Type 1 diabetes mellitus with diabetic amyotrophy |
| ICD-10-CM | E1049 | Type 1 diabetes mellitus with other diabetic neurological complication |
| ICD-10-CM | E1051 | Type 1 diabetes mellitus with diabetic peripheral angiopathy without gangrene |
| ICD-10-CM | E1052 | Type 1 diabetes mellitus with diabetic peripheral angiopathy with gangrene |
| ICD-10-CM | E1059 | Type 1 diabetes mellitus with other circulatory complications |
| ICD-10-CM | E10610 | Type 1 diabetes mellitus with diabetic neuropathic arthropathy |
| ICD-10-CM | E10618 | Type 1 diabetes mellitus with other diabetic arthropathy |
| ICD-10-CM | E10620 | Type 1 diabetes mellitus with diabetic dermatitis |
| ICD-10-CM | E10621 | Type 1 diabetes mellitus with foot ulcer |
| ICD-10-CM | E10622 | Type 1 diabetes mellitus with other skin ulcer |
| ICD-10-CM | E10628 | Type 1 diabetes mellitus with other skin complications |
| ICD-10-CM | E10630 | Type 1 diabetes mellitus with periodontal disease |
| ICD-10-CM | E10638 | Type 1 diabetes mellitus with other oral complications |
| ICD-10-CM | E10641 | Type 1 diabetes mellitus with hypoglycemia with coma |
| ICD-10-CM | E10649 | Type 1 diabetes mellitus with hypoglycemia without coma |
| ICD-10-CM | E1065 | Type 1 diabetes mellitus with hyperglycemia |
| ICD-10-CM | E1069 | Type 1 diabetes mellitus with other specified complication |
| ICD-10-CM | E108 | Type 1 diabetes mellitus with unspecified complications |
| ICD-10-CM | E109 | Type 1 diabetes mellitus without complications |
| ICD-10-CM | O24011 | Pre-existing type 1 diabetes mellitus, in pregnancy, first trimester |
| ICD-10-CM | O24012 | Pre-existing type 1 diabetes mellitus, in pregnancy, second trimester |

**Exhibit A4. ICD-10-CM Diagnosis Codes for Type 1 Diabetes**

| Code Type | Code | Description |
|---|---|---|
| ICD-10-CM | O24013 | Pre-existing type 1 diabetes mellitus, in pregnancy, third trimester |
| ICD-10-CM | O24019 | Pre-existing type 1 diabetes mellitus, in pregnancy, unspecified trimester |
| ICD-10-CM | O2402 | Pre-existing type 1 diabetes mellitus, in childbirth |
| ICD-10-CM | O2403 | Pre-existing type 1 diabetes mellitus, in the puerperium |

**Exhibit A5. NDC Codes for T2DM Prescription Drugs Used to Exclude Patients from the T1DM Population**

NDCs were generated for each of the drug, description, and use categories listed below. The full list of NDCs is available from the authors.
The use of GLP-1s to identify non-T1DM patients may need to be revisited if more recent data is analyzed,due to expanded GLP-1 indicaitons.

| Code Type | Drug Category | Description | Diabetes Patient Identification Use |
|---|---|---|---|
| NDC | AGI | Miglitol Tab 25 MG | Hypoglycemics/Antihyperglycemics |
| NDC | AGI | Miglitol Tab 50 MG | Hypoglycemics/Antihyperglycemics |
| NDC | AGI | Miglitol Tab 100 MG | Hypoglycemics/Antihyperglycemics |
| NDC | AGI | Acarbose Tab 50 MG | Hypoglycemics/Antihyperglycemics |
| NDC | AGI | Acarbose Tab 100 MG | Hypoglycemics/Antihyperglycemics |
| NDC | AGI | Acarbose Tab 25 MG | Hypoglycemics/Antihyperglycemics |
| NDC | DPP-4 | Saxagliptin HCl Tab 2.5 MG (Base Equiv) | Hypoglycemics/Antihyperglycemics |
| NDC | DPP-4 | Saxagliptin HCl Tab 5 MG (Base Equiv) | Hypoglycemics/Antihyperglycemics |
| NDC | DPP-4 | Sitagliptin Phosphate Tab 50 MG (Base Equiv) | Hypoglycemics/Antihyperglycemics |
| NDC | DPP-4 | Sitagliptin Phosphate Tab 25 MG (Base Equiv) | Hypoglycemics/Antihyperglycemics |
| NDC | DPP-4 | Sitagliptin Phosphate Tab 100 MG (Base Equiv) | Hypoglycemics/Antihyperglycemics |
| NDC | DPP-4 | Linagliptin Tab 5 MG | Hypoglycemics/Antihyperglycemics |
| NDC | DPP-4 | Alogliptin Benzoate Tab 6.25 MG (Base Equiv) | Hypoglycemics/Antihyperglycemics |
| NDC | DPP-4 | Alogliptin Benzoate Tab 12.5 MG (Base Equiv) | Hypoglycemics/Antihyperglycemics |
| NDC | DPP-4 | Alogliptin Benzoate Tab 25 MG (Base Equiv) | Hypoglycemics/Antihyperglycemics |
| NDC | DPP-4-Metformin | Saxagliptin-Metformin HCl Tab SR 24HR 5-500 MG | Hypoglycemics/Antihyperglycemics |
| NDC | DPP-4-Metformin | Saxagliptin-Metformin HCl Tab SR 24HR 2.5-1000 MG | Hypoglycemics/Antihyperglycemics |
| NDC | DPP-4-Metformin | Saxagliptin-Metformin HCl Tab SR 24HR 5-1000 MG | Hypoglycemics/Antihyperglycemics |
| NDC | DPP-4-Metformin | Sitagliptin-Metformin HCl Tab SR 24HR 50-500 MG | Hypoglycemics/Antihyperglycemics |
| NDC | DPP-4-Metformin | Sitagliptin-Metformin HCl Tab ER 24HR 50-1000 MG | Hypoglycemics/Antihyperglycemics |
| NDC | DPP-4-Metformin | Sitagliptin-Metformin HCl Tab SR 24HR 50-1000 MG | Hypoglycemics/Antihyperglycemics |
| NDC | DPP-4-Metformin | Sitagliptin-Metformin HCl Tab ER 24HR 100-1000 MG | Hypoglycemics/Antihyperglycemics |
| NDC | DPP-4-Metformin | Sitagliptin-Metformin HCl Tab SR 24HR 100-1000 MG | Hypoglycemics/Antihyperglycemics |
| NDC | DPP-4-Metformin | Sitagliptin-Metformin HCl Tab 50-500 MG | Hypoglycemics/Antihyperglycemics |
| NDC | DPP-4-Metformin | Sitagliptin-Metformin HCl Tab 50-1000 MG | Hypoglycemics/Antihyperglycemics |
| NDC | DPP-4-Metformin | Linagliptin-Metformin HCl Tab 2.5-500 MG | Hypoglycemics/Antihyperglycemics |
| NDC | DPP-4-Metformin | Linagliptin-Metformin HCl Tab 2.5-850 MG | Hypoglycemics/Antihyperglycemics |
| NDC | DPP-4-Metformin | Linagliptin-Metformin HCl Tab 2.5-1000 MG | Hypoglycemics/Antihyperglycemics |
| NDC | DPP-4-Metformin | Linagliptin-Metformin HCl Tab SR 24HR 2.5-1000 MG | Hypoglycemics/Antihyperglycemics |
| NDC | DPP-4-Metformin | Linagliptin-Metformin HCl Tab SR 24HR 5-1000 MG | Hypoglycemics/Antihyperglycemics |
| NDC | DPP-4-Metformin | Alogliptin-Metformin HCl Tab 12.5-500 MG | Hypoglycemics/Antihyperglycemics |
| NDC | DPP-4-Metformin | Alogliptin-Metformin HCl Tab 12.5-1000 MG | Hypoglycemics/Antihyperglycemics |
| NDC | DPP-4-Thiazolidinedione | Alogliptin-Pioglitazone Tab 12.5-15 MG | Hypoglycemics/Antihyperglycemics |
| NDC | DPP-4-Thiazolidinedione | Alogliptin-Pioglitazone Tab 12.5-30 MG | Hypoglycemics/Antihyperglycemics |
| NDC | DPP-4-Thiazolidinedione | Alogliptin-Pioglitazone Tab 12.5-45 MG | Hypoglycemics/Antihyperglycemics |
| NDC | DPP-4-Thiazolidinedione | Alogliptin-Pioglitazone Tab 25-15 MG | Hypoglycemics/Antihyperglycemics |
| NDC | DPP-4-Thiazolidinedione | Alogliptin-Pioglitazone Tab 25-30 MG | Hypoglycemics/Antihyperglycemics |
| NDC | DPP-4-Thiazolidinedione | Alogliptin-Pioglitazone Tab 25-45 MG | Hypoglycemics/Antihyperglycemics |
| NDC | DPP-4-HMG-CoA Inhibitor | Sitagliptin-Simvastatin Tab 50-10 MG | Hypoglycemics/Antihyperglycemics |
| NDC | DPP-4-HMG-CoA Inhibitor | Sitagliptin-Simvastatin Tab 50-20 MG | Hypoglycemics/Antihyperglycemics |
| NDC | DPP-4-HMG-CoA Inhibitor | Sitagliptin-Simvastatin Tab 50-40 MG | Hypoglycemics/Antihyperglycemics |
| NDC | DPP-4-HMG-CoA Inhibitor | Sitagliptin-Simvastatin Tab 100-10 MG | Hypoglycemics/Antihyperglycemics |
| NDC | DPP-4-HMG-CoA Inhibitor | Sitagliptin-Simvastatin Tab 100-20 MG | Hypoglycemics/Antihyperglycemics |
| NDC | DPP-4-HMG-CoA Inhibitor | Sitagliptin-Simvastatin Tab 100-40 MG | Hypoglycemics/Antihyperglycemics |
| NDC | Glinide | Nateglinide Tab 60 MG | Hypoglycemics/Antihyperglycemics |
| NDC | Glinide | Nateglinide Tab 120 MG | Hypoglycemics/Antihyperglycemics |
| NDC | Glinide | Repaglinide Tab 0.5 MG | Hypoglycemics/Antihyperglycemics |
| NDC | Glinide | Repaglinide Tab 1 MG | Hypoglycemics/Antihyperglycemics |
| NDC | Glinide | Repaglinide Tab 2 MG | Hypoglycemics/Antihyperglycemics |
| NDC | Glinide-Metformin | Repaglinide-Metformin HCl Tab 2-500 MG | Hypoglycemics/Antihyperglycemics |
| NDC | Glinide-Metformin | Repaglinide-Metformin HCl Tab 1-500 MG | Hypoglycemics/Antihyperglycemics |
| NDC | GLP-1 | Dulaglutide Soln Pen-injector 0.75 MG/0.5ML | Hypoglycemics/Antihyperglycemics |
| NDC | GLP-1 | Dulaglutide Soln Pen-injector 1.5 MG/0.5ML | Hypoglycemics/Antihyperglycemics |
| NDC | GLP-1 | Dulaglutide Soln Pen-injector 3 MG/0.5ML | Hypoglycemics/Antihyperglycemics |
| NDC | GLP-1 | Dulaglutide Soln Pen-injector 4.5 MG/0.5ML | Hypoglycemics/Antihyperglycemics |
| NDC | GLP-1 | Lixisenatide Soln Pen-injector 20 MCG/0.2ML (100 MCG/ML) | Hypoglycemics/Antihyperglycemics |
| NDC | GLP-1 | Lixisenatide Pen-inj Starter Kit 10 MCG/0.2ML & 20 MCG/0.2ML | Hypoglycemics/Antihyperglycemics |
| NDC | GLP-1 | Liraglutide (Weight Mngmt) Soln Pen-Inj 18 MG/3ML (6 MG/ML) | Hypoglycemics/Antihyperglycemics |
| NDC | GLP-1 | Liraglutide Soln Pen-injector 18 MG/3ML (6 MG/ML) | Hypoglycemics/Antihyperglycemics |
| NDC | GLP-1 | Semaglutide Soln Pen-inj 0.25 or 0.5 MG/DOSE (2 MG/1.5ML) | Hypoglycemics/Antihyperglycemics |
| NDC | GLP-1 | Semaglutide Soln Pen-inj 1 MG/DOSE (2 MG/1.5ML) | Hypoglycemics/Antihyperglycemics |
| NDC | GLP-1 | Semaglutide Tab 3 MG | Hypoglycemics/Antihyperglycemics |
| NDC | GLP-1 | Semaglutide Tab 7 MG | Hypoglycemics/Antihyperglycemics |
| NDC | GLP-1 | Semaglutide Tab 14 MG | Hypoglycemics/Antihyperglycemics |
| NDC | GLP-1 | Albiglutide For Soln Pen-injector 30 MG | Hypoglycemics/Antihyperglycemics |
| NDC | GLP-1 | Albiglutide For Soln Pen-injector 50 MG | Hypoglycemics/Antihyperglycemics |
| NDC | GLP-1 | Exenatide Soln Pen-injector 5 MCG/0.02ML | Hypoglycemics/Antihyperglycemics |
| NDC | GLP-1 | Exenatide Extended Release for Inj Susp 2 MG | Hypoglycemics/Antihyperglycemics |
| NDC | GLP-1 | Exenatide Soln Pen-injector 10 MCG/0.04ML | Hypoglycemics/Antihyperglycemics |
| NDC | GLP-1 | Exenatide Extended Release for Susp Pen-injector 2 MG | Hypoglycemics/Antihyperglycemics |
| NDC | GLP-1 | Exenatide Extended Release Susp Auto-Injector 2 MG/0.85ML | Hypoglycemics/Antihyperglycemics |
| NDC | GLP-1 | Exenatide Inj 10 MCG/0.04ML | Hypoglycemics/Antihyperglycemics |
| NDC | GLP-1 | Exenatide 2 mg subcutaneous injection, extended release | Hypoglycemics/Antihyperglycemics |
| NDC | SGLT2 | Dapagliflozin Propanediol Tab 5 MG (Base Equivalent) | Hypoglycemics/Antihyperglycemics |
| NDC | SGLT2 | Dapagliflozin Propanediol Tab 10 MG (Base Equivalent) | Hypoglycemics/Antihyperglycemics |
| NDC | SGLT2 | Ertugliflozin L-Pyroglutamic Acid Tab 5 MG (Base Equiv) | Hypoglycemics/Antihyperglycemics |
| NDC | SGLT2 | Ertugliflozin L-Pyroglutamic Acid Tab 15 MG (Base Equiv) | Hypoglycemics/Antihyperglycemics |

**Exhibit A5. NDC Codes for T2DM Prescription Drugs Used to Exclude Patients from the T1DM Population**

NDCs were generated for each of the drug, description, and use categories listed below. The full list of NDCs is available from the authors.
The use of GLP-1s to identify non-T1DM patients may need to be revisited if more recent data is analyzed,due to expanded GLP-1 indicaitons.

| Code Type | Drug Category | Description | Diabetes Patient Identification Use |
|---|---|---|---|
| NDC | AGI | Miglitol Tab 25 MG | Hypoglycemics/Antihyperglycemics |
| NDC | SGLT2 | Empagliflozin Tab 10 MG | Hypoglycemics/Antihyperglycemics |
| NDC | SGLT2 | Empagliflozin Tab 25 MG | Hypoglycemics/Antihyperglycemics |
| NDC | SGLT2 | Canagliflozin Tab 100 MG | Hypoglycemics/Antihyperglycemics |
| NDC | SGLT2 | Canagliflozin Tab 300 MG | Hypoglycemics/Antihyperglycemics |
| NDC | SGLT2-DPP-4 | Ertugliflozin-Sitagliptin Tab 5-100 MG | Hypoglycemics/Antihyperglycemics |
| NDC | SGLT2-DPP-4 | Ertugliflozin-Sitagliptin Tab 15-100 MG | Hypoglycemics/Antihyperglycemics |
| NDC | SGLT2-DPP-4 | Dapagliflozin-Saxagliptin Tab 5-5 MG | Hypoglycemics/Antihyperglycemics |
| NDC | SGLT2-DPP-4 | Dapagliflozin-Saxagliptin Tab 10-5 MG | Hypoglycemics/Antihyperglycemics |
| NDC | SGLT2-DPP-4 | Empagliflozin-Linagliptin Tab 25-5 MG | Hypoglycemics/Antihyperglycemics |
| NDC | SGLT2-DPP-4 | Empagliflozin-Linagliptin Tab 10-5 MG | Hypoglycemics/Antihyperglycemics |
| NDC | SGLT2-Metformin | Ertugliflozin-Metformin HCl Tab 2.5-500 MG | Hypoglycemics/Antihyperglycemics |
| NDC | SGLT2-Metformin | Ertugliflozin-Metformin HCl Tab 7.5-500 MG | Hypoglycemics/Antihyperglycemics |
| NDC | SGLT2-Metformin | Ertugliflozin-Metformin HCl Tab 2.5-1000 MG | Hypoglycemics/Antihyperglycemics |
| NDC | SGLT2-Metformin | Ertugliflozin-Metformin HCl Tab 7.5-1000 MG | Hypoglycemics/Antihyperglycemics |
| NDC | SGLT2-Metformin | Dapagliflozin-Metformin HCl Tab ER 24HR 2.5-1000 MG | Hypoglycemics/Antihyperglycemics |
| NDC | SGLT2-Metformin | Dapagliflozin-Metformin HCl Tab SR 24HR 5-500 MG | Hypoglycemics/Antihyperglycemics |
| NDC | SGLT2-Metformin | Dapagliflozin-Metformin HCl Tab SR 24HR 5-1000 MG | Hypoglycemics/Antihyperglycemics |
| NDC | SGLT2-Metformin | Dapagliflozin-Metformin HCl Tab SR 24HR 10-500 MG | Hypoglycemics/Antihyperglycemics |
| NDC | SGLT2-Metformin | Dapagliflozin-Metformin HCl Tab SR 24HR 10-1000 MG | Hypoglycemics/Antihyperglycemics |
| NDC | SGLT2-Metformin | Empagliflozin-Metformin HCl Tab 5-500 MG | Hypoglycemics/Antihyperglycemics |
| NDC | SGLT2-Metformin | Empagliflozin-Metformin HCl Tab 12.5-1000 MG | Hypoglycemics/Antihyperglycemics |
| NDC | SGLT2-Metformin | Empagliflozin-Metformin HCl Tab 5-1000 MG | Hypoglycemics/Antihyperglycemics |
| NDC | SGLT2-Metformin | Empagliflozin-Metformin HCl Tab 12.5-500 MG | Hypoglycemics/Antihyperglycemics |
| NDC | SGLT2-Metformin | Empagliflozin-Metformin HCl Tab SR 24HR 10-1000 MG | Hypoglycemics/Antihyperglycemics |
| NDC | SGLT2-Metformin | Empagliflozin-Metformin HCl Tab SR 24HR 5-1000 MG | Hypoglycemics/Antihyperglycemics |
| NDC | SGLT2-Metformin | Empagliflozin-Metformin HCl Tab SR 24HR 25-1000 MG | Hypoglycemics/Antihyperglycemics |
| NDC | SGLT2-Metformin | Empagliflozin-Metformin HCl Tab SR 24HR 12.5-1000 MG | Hypoglycemics/Antihyperglycemics |
| NDC | SGLT2-Metformin | Canagliflozin-Metformin HCl Tab 50-500 MG | Hypoglycemics/Antihyperglycemics |
| NDC | SGLT2-Metformin | Canagliflozin-Metformin HCl Tab 50-1000 MG | Hypoglycemics/Antihyperglycemics |
| NDC | SGLT2-Metformin | Canagliflozin-Metformin HCl Tab 150-500 MG | Hypoglycemics/Antihyperglycemics |
| NDC | SGLT2-Metformin | Canagliflozin-Metformin HCl Tab 150-1000 MG | Hypoglycemics/Antihyperglycemics |
| NDC | SGLT2-Metformin | Canagliflozin-Metformin HCl Tab SR 24HR 50-500 MG | Hypoglycemics/Antihyperglycemics |
| NDC | SGLT2-Metformin | Canagliflozin-Metformin HCl Tab SR 24HR 50-1000 MG | Hypoglycemics/Antihyperglycemics |
| NDC | SGLT2-Metformin | Canagliflozin-Metformin HCl Tab SR 24HR 150-500 MG | Hypoglycemics/Antihyperglycemics |
| NDC | SGLT2-Metformin | Canagliflozin-Metformin HCl Tab SR 24HR 150-1000 MG | Hypoglycemics/Antihyperglycemics |
| NDC | Sulfonylurea | Glyburide Micronized Tab 1.5 MG | Hypoglycemics/Antihyperglycemics |
| NDC | Sulfonylurea | Glyburide Micronized Tab 3 MG | Hypoglycemics/Antihyperglycemics |
| NDC | Sulfonylurea | Glyburide Micronized Tab 6 MG | Hypoglycemics/Antihyperglycemics |
| NDC | Sulfonylurea | Glyburide Tab 2.5 MG | Hypoglycemics/Antihyperglycemics |
| NDC | Sulfonylurea | Glyburide Tab 5 MG | Hypoglycemics/Antihyperglycemics |
| NDC | Sulfonylurea | Glyburide Tab 1.25 MG | Hypoglycemics/Antihyperglycemics |
| NDC | Sulfonylurea | Glimepiride Tab 1 MG | Hypoglycemics/Antihyperglycemics |
| NDC | Sulfonylurea | Glimepiride Tab 2 MG | Hypoglycemics/Antihyperglycemics |
| NDC | Sulfonylurea | Glimepiride Tab 4 MG | Hypoglycemics/Antihyperglycemics |
| NDC | Sulfonylurea | Glipizide Tab SR 24HR 2.5 MG | Hypoglycemics/Antihyperglycemics |
| NDC | Sulfonylurea | Glipizide Tab SR 24HR 5 MG | Hypoglycemics/Antihyperglycemics |
| NDC | Sulfonylurea | Glipizide Tab SR 24HR 10 MG | Hypoglycemics/Antihyperglycemics |
| NDC | Sulfonylurea | Glipizide Tab 5 MG | Hypoglycemics/Antihyperglycemics |
| NDC | Sulfonylurea | Glipizide Tab 10 MG | Hypoglycemics/Antihyperglycemics |
| NDC | Sulfonylurea | Glyburide 2.5 mg oral tablet | Hypoglycemics/Antihyperglycemics |
| NDC | Sulfonylurea | Glipizide 5 mg oral tablet | Hypoglycemics/Antihyperglycemics |
| NDC | Sulfonylurea | Chlorpropamide Tab 250 MG | Hypoglycemics/Antihyperglycemics |
| NDC | Sulfonylurea | Glyburide 5 mg oral tablet | Hypoglycemics/Antihyperglycemics |
| NDC | Sulfonylurea | Glipizide 10 mg oral tablet | Hypoglycemics/Antihyperglycemics |
| NDC | Sulfonylurea | Glipizide 10 mg oral tablet, extended release | Hypoglycemics/Antihyperglycemics |
| NDC | Sulfonylurea | Glipizide 5 mg oral tablet, extended release | Hypoglycemics/Antihyperglycemics |
| NDC | Sulfonylurea | Glyburide 1.25 mg oral tablet | Hypoglycemics/Antihyperglycemics |
| NDC | Sulfonylurea | Glimepiride 1 mg oral tablet | Hypoglycemics/Antihyperglycemics |
| NDC | Sulfonylurea | Glimepiride 2 mg oral tablet | Hypoglycemics/Antihyperglycemics |
| NDC | Sulfonylurea | Chlorpropamide Tab 100 MG | Hypoglycemics/Antihyperglycemics |
| NDC | Sulfonylurea | Tolbutamide Tab 500 MG | Hypoglycemics/Antihyperglycemics |
| NDC | Sulfonylurea | Tolazamide Tab 250 MG | Hypoglycemics/Antihyperglycemics |
| NDC | Sulfonylurea | Tolazamide Tab 500 MG | Hypoglycemics/Antihyperglycemics |
| NDC | Sulfonylurea | Glipizide Tab ER 24HR 2.5 MG | Hypoglycemics/Antihyperglycemics |
| NDC | Sulfonylurea | Glipizide Tab ER 24HR 5 MG | Hypoglycemics/Antihyperglycemics |
| NDC | Sulfonylurea | Glipizide Tab ER 24HR 10 MG | Hypoglycemics/Antihyperglycemics |
| NDC | Sulfonylurea | Glipizide Powder | Hypoglycemics/Antihyperglycemics |
| NDC | Sulfonylurea | Glyburide Powder | Hypoglycemics/Antihyperglycemics |
| NDC | Sulfonylurea | Chlorpropamid 250 mg oral tablet | Hypoglycemics/Antihyperglycemics |
| NDC | Sulfonylurea | Glimepiride 4 mg oral tablet | Hypoglycemics/Antihyperglycemics |
| NDC | Sulfonylurea | Glyburide Micronized Tab 4.5 MG | Hypoglycemics/Antihyperglycemics |
| NDC | Sulfonylurea | Glyburide micronized 1.5 mg oral tablet | Hypoglycemics/Antihyperglycemics |
| NDC | Sulfonylurea | Glyburide Tab 1.25 MG | Hypoglycemics/Antihyperglycemics |
| NDC | Sulfonylurea-Metformin | Glyburide-Metformin Tab 1.25-250 MG | Hypoglycemics/Antihyperglycemics |
| NDC | Sulfonylurea-Metformin | Glyburide-Metformin Tab 2.5-500 MG | Hypoglycemics/Antihyperglycemics |
| NDC | Sulfonylurea-Metformin | Glyburide-Metformin Tab 5-500 MG | Hypoglycemics/Antihyperglycemics |
| NDC | Sulfonylurea-Metformin | Glipizide-Metformin HCl Tab 2.5-500 MG | Hypoglycemics/Antihyperglycemics |

**Exhibit A5. NDC Codes for T2DM Prescription Drugs Used to Exclude Patients from the T1DM Population**

NDCs were generated for each of the drug, description, and use categories listed below. The full list of NDCs is available from the authors.
The use of GLP-1s to identify non-T1DM patients may need to be revisited if more recent data is analyzed,due to expanded GLP-1 indicaitons.

| Code Type | Drug Category | Description | Diabetes Patient Identification Use |
|---|---|---|---|
| NDC | AGI | Miglitol Tab 25 MG | Hypoglycemics/Antihyperglycemics |
| NDC | Sulfonylurea-Metformin | Glipizide-Metformin HCl Tab 5-500 MG | Hypoglycemics/Antihyperglycemics |
| NDC | Sulfonylurea-Metformin | Glipizide-Metformin HCl Tab 2.5-250 MG | Hypoglycemics/Antihyperglycemics |
| NDC | Sulfonylurea-Metformin | Glyburide-metformin 1.25 mg-250 mg oral tablet | Hypoglycemics/Antihyperglycemics |
| NDC | Sulfonylurea-Metformin | Glyburide-metformin 2.5 mg-500 mg oral tablet | Hypoglycemics/Antihyperglycemics |
| NDC | Sulfonylurea-Metformin | Glyburide-metformin 5 mg-500 mg oral tablet | Hypoglycemics/Antihyperglycemics |
| NDC | Sulfonylurea-Metformin | Glyburide Micronized Tab 1.5 MG | Hypoglycemics/Antihyperglycemics |
| NDC | Sulfonylurea-Thiazolidinedione | Rosiglitazone Maleate-Glimepiride Tab 8-2 MG | Hypoglycemics/Antihyperglycemics |
| NDC | Sulfonylurea-Thiazolidinedione | Rosiglitazone Maleate-Glimepiride Tab 8-4 MG | Hypoglycemics/Antihyperglycemics |
| NDC | Sulfonylurea-Thiazolidinedione | Rosiglitazone Maleate-Glimepiride Tab 4-1 MG | Hypoglycemics/Antihyperglycemics |
| NDC | Sulfonylurea-Thiazolidinedione | Rosiglitazone Maleate-Glimepiride Tab 4-2 MG | Hypoglycemics/Antihyperglycemics |
| NDC | Sulfonylurea-Thiazolidinedione | Rosiglitazone Maleate-Glimepiride Tab 4-4 MG | Hypoglycemics/Antihyperglycemics |
| NDC | Sulfonylurea-Thiazolidinedione | Pioglitazone HCl-Glimepiride Tab 30-2 MG | Hypoglycemics/Antihyperglycemics |
| NDC | Sulfonylurea-Thiazolidinedione | Pioglitazone HCl-Glimepiride Tab 30-4 MG | Hypoglycemics/Antihyperglycemics |
| NDC | Thiazolidinedione | Rosiglitazone Maleate Tab 2 MG (Base Equiv) | Hypoglycemics/Antihyperglycemics |
| NDC | Thiazolidinedione | Rosiglitazone Maleate Tab 4 MG (Base Equiv) | Hypoglycemics/Antihyperglycemics |
| NDC | Thiazolidinedione | Rosiglitazone Maleate Tab 8 MG (Base Equiv) | Hypoglycemics/Antihyperglycemics |
| NDC | Thiazolidinedione | Pioglitazone HCl Tab 45 MG (Base Equiv) | Hypoglycemics/Antihyperglycemics |
| NDC | Thiazolidinedione | Pioglitazone HCl Tab 30 MG (Base Equiv) | Hypoglycemics/Antihyperglycemics |
| NDC | Thiazolidinedione | Pioglitazone HCl Tab 15 MG (Base Equiv) | Hypoglycemics/Antihyperglycemics |
| NDC | Thiazolidinedione | Pioglitazone 45 mg oral tablet | Hypoglycemics/Antihyperglycemics |
| NDC | Thiazolidinedione | Rosiglitazone 4 mg oral tablet | Hypoglycemics/Antihyperglycemics |
| NDC | Thiazolidinedione | Rosiglitazone 8 mg oral tablet | Hypoglycemics/Antihyperglycemics |
| NDC | Thiazolidinedione | Pioglitazone 30 mg oral tablet | Hypoglycemics/Antihyperglycemics |
| NDC | Thiazolidinedione | Pioglitazone 15 mg oral tablet | Hypoglycemics/Antihyperglycemics |
| NDC | Thiazolidinedione | Rosiglitazone Maleate-Metformin HCl Tab 2-500 MG | Hypoglycemics/Antihyperglycemics |
| NDC | Thiazolidinedione-Metformin | Rosiglitazone Maleate-Metformin HCl Tab 2-1000 MG | Hypoglycemics/Antihyperglycemics |
| NDC | Thiazolidinedione-Metformin | Rosiglitazone Maleate-Metformin HCl Tab 4-1000 MG | Hypoglycemics/Antihyperglycemics |
| NDC | Thiazolidinedione-Metformin | Rosiglitazone Maleate-Metformin HCl Tab 2-500 MG | Hypoglycemics/Antihyperglycemics |
| NDC | Thiazolidinedione-Metformin | Rosiglitazone Maleate-Metformin HCl Tab 4-500 MG | Hypoglycemics/Antihyperglycemics |
| NDC | Thiazolidinedione-Metformin | Pioglitazone HCl-Metformin HCl Tab 15-500 MG | Hypoglycemics/Antihyperglycemics |
| NDC | Thiazolidinedione-Metformin | Pioglitazone HCl-Metformin HCl Tab 15-850 MG | Hypoglycemics/Antihyperglycemics |
| NDC | Thiazolidinedione-Metformin | Pioglitazone HCl-Metformin HCl Tab SR 24HR 30-1000 MG | Hypoglycemics/Antihyperglycemics |
| NDC | Thiazolidinedione-Metformin | Pioglitazone HCl-Metformin HCl Tab SR 24HR 15-1000 MG | Hypoglycemics/Antihyperglycemics |

**Exhibit A6. State Region Mapping**

| State | Mapped Region |
|---|---|
| Alaska | West |
| Alabama | South |
| Arkansas | South |
| Arizona | West |
| California | West |
| Colorado | West |
| Connecticut | Northeast |
| District of Columbia | South |
| Delaware | South |
| Florida | South |
| Georgia | South |
| Hawaii | West |
| Iowa | Midwest |
| Idaho | West |
| Illinois | Midwest |
| Indiana | Midwest |
| Kansas | Midwest |
| Kentucky | South |
| Louisiana | South |
| Massachusetts | Northeast |
| Maryland | South |
| Maine | Northeast |
| Michigan | Midwest |
| Minnesota | Midwest |
| Missouri | Midwest |
| Mississippi | South |
| Montana | West |
| North Carolina | South |
| North Dakota | Midwest |
| Nebraska | Midwest |
| New Hampshire | Northeast |
| New Jersey | Northeast |
| New Mexico | West |
| Nevada | West |
| New York | Northeast |
| Ohio | Midwest |
| Oklahoma | South |
| Oregon | West |
| Pennsylvania | Northeast |
| Rhode Island | Northeast |
| South Carolina | South |
| South Dakota | Midwest |
| Tennessee | South |
| Texas | South |
| Utah | West |
| Virginia | South |
| Vermont | Northeast |
| Washington | West |
| Wisconsin | Midwest |
| West Virginia | South |
| Wyoming | West |

**Exhibit A7. Prevalence and Extrapolation - Total T1DM Population**

| Description | Source | FFS (a AND/OR b) | MA-PD (c AND/OR D) | Other Medicare Health Plan Enrollment | Commercial (<65) | Medicaid (excluding duals) | VA (18-64) | Uninsured (<65)* |
|---|---|---|---|---|---|---|---|---|
| 2019 National Enrollment | Literature Review | 38,577,012 | 22,200,000 | 3,622,988 | 172,700,000 | 64,896,849 | 4,236,144 | 32,800,000 |
| Annual Prevalence | Claims Analysis | 0.867% | 0.863% | 0.867% | 0.488% | 0.406% | 0.488% | 0.406% |
| Extrapolated to 2019 | 2019 National Enrollment * Annual Prevalence | 334,524 | 191,498 | 31,417 | 843,567 | 263,710 | 20,692 | 133,283 |

**Exhibit A8. Standard Mortality Rates by Age and Sex**

CDC WONDER. *Multiple Cause of Death, 1999-2020 Request* .
https://wonder.cdc.gov/mcd.html

| Age | M/F | WONDER Population | WONDER Deaths | WONDER Mortality Rate |
|---|---|---|---|---|
| 0 | M | 1,935,117 | 11,674 | 0.603% |
| 1 | M | 1,958,585 | 825 | 0.042% |
| 2 | M | 2,005,544 | 515 | 0.026% |
| 3 | M | 2,043,010 | 390 | 0.019% |
| 4 | M | 2,066,951 | 311 | 0.015% |
| 5 | M | 2,061,200 | 291 | 0.014% |
| 6 | M | 2,052,956 | 266 | 0.013% |
| 7 | M | 2,055,735 | 243 | 0.012% |
| 8 | M | 2,079,723 | 251 | 0.012% |
| 9 | M | 2,073,148 | 239 | 0.012% |
| 10 | M | 2,071,778 | 291 | 0.014% |
| 11 | M | 2,138,037 | 324 | 0.015% |
| 12 | M | 2,149,819 | 351 | 0.016% |
| 13 | M | 2,132,987 | 394 | 0.018% |
| 14 | M | 2,125,640 | 536 | 0.025% |
| 15 | M | 2,129,720 | 754 | 0.035% |
| 16 | M | 2,116,165 | 1,040 | 0.049% |
| 17 | M | 2,112,553 | 1,353 | 0.064% |
| 18 | M | 2,172,385 | 1,972 | 0.091% |
| 19 | M | 2,214,784 | 2,242 | 0.101% |
| 20 | M | 2,183,139 | 2,452 | 0.112% |
| 21 | M | 2,187,086 | 2,871 | 0.131% |
| 22 | M | 2,195,296 | 2,909 | 0.133% |
| 23 | M | 2,221,758 | 2,979 | 0.134% |
| 24 | M | 2,277,473 | 3,176 | 0.139% |
| 25 | M | 2,325,853 | 3,481 | 0.150% |
| 26 | M | 2,365,886 | 3,696 | 0.156% |
| 27 | M | 2,421,270 | 3,864 | 0.160% |
| 28 | M | 2,451,756 | 4,129 | 0.168% |
| 29 | M | 2,439,805 | 4,272 | 0.175% |
| 30 | M | 2,340,865 | 4,301 | 0.184% |
| 31 | M | 2,281,327 | 4,282 | 0.188% |
| 32 | M | 2,239,886 | 4,269 | 0.191% |
| 33 | M | 2,243,280 | 4,410 | 0.197% |
| 34 | M | 2,249,252 | 4,647 | 0.207% |
| 35 | M | 2,167,622 | 4,671 | 0.215% |
| 36 | M | 2,191,368 | 4,854 | 0.222% |
| 37 | M | 2,181,162 | 5,066 | 0.232% |
| 38 | M | 2,149,359 | 5,281 | 0.246% |
| 39 | M | 2,195,430 | 5,369 | 0.245% |
| 40 | M | 2,046,663 | 5,628 | 0.275% |
| 41 | M | 2,001,502 | 5,385 | 0.269% |
| 42 | M | 1,972,584 | 5,655 | 0.287% |
| 43 | M | 1,913,289 | 5,558 | 0.290% |
| 44 | M | 1,973,101 | 5,969 | 0.303% |
| 45 | M | 1,900,492 | 6,309 | 0.332% |
| 46 | M | 1,919,298 | 6,937 | 0.361% |
| 47 | M | 2,005,221 | 7,793 | 0.389% |
| 48 | M | 2,114,614 | 8,843 | 0.418% |
| 49 | M | 2,145,730 | 9,476 | 0.442% |
| 50 | M | 2,025,376 | 9,940 | 0.491% |
| 51 | M | 1,976,883 | 10,653 | 0.539% |
| 52 | M | 1,968,619 | 11,605 | 0.589% |
| 53 | M | 2,001,325 | 12,550 | 0.627% |
| 54 | M | 2,114,408 | 14,741 | 0.697% |
| 55 | M | 2,137,586 | 16,534 | 0.773% |
| 56 | M | 2,125,123 | 17,793 | 0.837% |
| 57 | M | 2,111,795 | 19,341 | 0.916% |
| 58 | M | 2,125,774 | 20,908 | 0.984% |
| 59 | M | 2,142,211 | 22,479 | 1.049% |
| 60 | M | 2,052,677 | 23,299 | 1.135% |

**Exhibit A8. Standard Mortality Rates by Age and Sex**

CDC WONDER. *Multiple Cause of Death, 1999-2020 Request* .
https://wonder.cdc.gov/mcd.html

| Age | M/F | WONDER Population | WONDER Deaths | WONDER Mortality Rate |
|---|---|---|---|---|
| 61 | M | 2,027,951 | 25,181 | 1.242% |
| 62 | M | 1,991,220 | 26,654 | 1.339% |
| 63 | M | 1,905,842 | 27,524 | 1.444% |
| 64 | M | 1,879,040 | 28,212 | 1.501% |
| 65 | M | 1,783,428 | 29,113 | 1.632% |
| 66 | M | 1,703,888 | 29,767 | 1.747% |
| 67 | M | 1,627,863 | 30,240 | 1.858% |
| 68 | M | 1,564,129 | 30,695 | 1.962% |
| 69 | M | 1,520,465 | 31,123 | 2.047% |
| 70 | M | 1,462,120 | 32,921 | 2.252% |
| 71 | M | 1,435,378 | 34,789 | 2.424% |
| 72 | M | 1,483,236 | 37,706 | 2.542% |
| 73 | M | 1,074,771 | 31,566 | 2.937% |
| 74 | M | 1,044,301 | 32,327 | 3.096% |
| 75 | M | 998,390 | 34,576 | 3.463% |
| 76 | M | 1,003,690 | 37,415 | 3.728% |
| 77 | M | 854,930 | 35,401 | 4.141% |
| 78 | M | 760,170 | 34,513 | 4.540% |
| 79 | M | 701,319 | 34,591 | 4.932% |
| 80 | M | 643,638 | 35,264 | 5.479% |
| 81 | M | 593,628 | 35,972 | 6.060% |
| 82 | M | 525,766 | 35,419 | 6.737% |
| 83 | M | 480,165 | 35,821 | 7.460% |
| 84 | M | 436,527 | 36,128 | 8.276% |
| 85+ | M | 2,376,488 | 338,165 | 14.230% |
| 0 | F | 1,847,935 | 9,247 | 0.500% |
| 1 | F | 1,871,014 | 649 | 0.035% |
| 2 | F | 1,916,500 | 401 | 0.021% |
| 3 | F | 1,955,655 | 320 | 0.016% |
| 4 | F | 1,976,372 | 265 | 0.013% |
| 5 | F | 1,967,081 | 245 | 0.012% |
| 6 | F | 1,964,271 | 224 | 0.011% |
| 7 | F | 1,966,584 | 196 | 0.010% |
| 8 | F | 1,986,471 | 184 | 0.009% |
| 9 | F | 1,988,726 | 194 | 0.010% |
| 10 | F | 1,989,162 | 189 | 0.010% |
| 11 | F | 2,051,224 | 212 | 0.010% |
| 12 | F | 2,058,568 | 250 | 0.012% |
| 13 | F | 2,042,234 | 293 | 0.014% |
| 14 | F | 2,038,819 | 324 | 0.016% |
| 15 | F | 2,045,739 | 383 | 0.019% |
| 16 | F | 2,034,255 | 445 | 0.022% |
| 17 | F | 2,029,872 | 533 | 0.026% |
| 18 | F | 2,083,442 | 760 | 0.036% |
| 19 | F | 2,115,655 | 776 | 0.037% |
| 20 | F | 2,086,544 | 861 | 0.041% |
| 21 | F | 2,091,237 | 950 | 0.045% |
| 22 | F | 2,103,476 | 973 | 0.046% |
| 23 | F | 2,119,886 | 1,167 | 0.055% |
| 24 | F | 2,167,045 | 1,175 | 0.054% |
| 25 | F | 2,213,205 | 1,271 | 0.057% |
| 26 | F | 2,245,334 | 1,390 | 0.062% |
| 27 | F | 2,312,599 | 1,519 | 0.066% |
| 28 | F | 2,366,969 | 1,628 | 0.069% |
| 29 | F | 2,366,339 | 1,719 | 0.073% |
| 30 | F | 2,273,519 | 1,882 | 0.083% |
| 31 | F | 2,220,984 | 1,943 | 0.087% |
| 32 | F | 2,181,619 | 1,994 | 0.091% |
| 33 | F | 2,189,693 | 2,273 | 0.104% |
| 34 | F | 2,210,880 | 2,208 | 0.100% |
| 35 | F | 2,148,244 | 2,327 | 0.108% |
| 36 | F | 2,181,076 | 2,472 | 0.113% |

**Exhibit A8. Standard Mortality Rates by Age and Sex**

CDC WONDER. *Multiple Cause of Death, 1999-2020 Request* .
https://wonder.cdc.gov/mcd.html

| Age | M/F | WONDER Population | WONDER Deaths | WONDER Mortality Rate |
|---|---|---|---|---|
| 37 | F | 2,180,124 | 2,626 | 0.120% |
| 38 | F | 2,156,217 | 2,896 | 0.134% |
| 39 | F | 2,186,919 | 3,076 | 0.141% |
| 40 | F | 2,058,650 | 3,011 | 0.146% |
| 41 | F | 2,018,752 | 3,071 | 0.152% |
| 42 | F | 2,002,157 | 3,136 | 0.157% |
| 43 | F | 1,940,751 | 3,370 | 0.174% |
| 44 | F | 1,994,174 | 3,565 | 0.179% |
| 45 | F | 1,937,417 | 3,861 | 0.199% |
| 46 | F | 1,970,074 | 4,261 | 0.216% |
| 47 | F | 2,052,817 | 4,786 | 0.233% |
| 48 | F | 2,168,043 | 5,542 | 0.256% |
| 49 | F | 2,184,045 | 5,931 | 0.272% |
| 50 | F | 2,071,196 | 6,211 | 0.300% |
| 51 | F | 2,027,460 | 6,545 | 0.323% |
| 52 | F | 2,033,163 | 7,183 | 0.353% |
| 53 | F | 2,067,526 | 8,043 | 0.389% |
| 54 | F | 2,191,195 | 9,183 | 0.419% |
| 55 | F | 2,236,979 | 10,259 | 0.459% |
| 56 | F | 2,235,893 | 11,173 | 0.500% |
| 57 | F | 2,230,590 | 12,317 | 0.552% |
| 58 | F | 2,259,796 | 13,439 | 0.595% |
| 59 | F | 2,271,644 | 14,210 | 0.626% |
| 60 | F | 2,199,986 | 15,391 | 0.700% |
| 61 | F | 2,187,221 | 16,341 | 0.747% |
| 62 | F | 2,165,425 | 17,321 | 0.800% |
| 63 | F | 2,090,246 | 17,837 | 0.853% |
| 64 | F | 2,071,538 | 18,724 | 0.904% |
| 65 | F | 1,991,169 | 19,046 | 0.957% |
| 66 | F | 1,914,181 | 19,793 | 1.034% |
| 67 | F | 1,836,574 | 20,479 | 1.115% |
| 68 | F | 1,781,346 | 21,517 | 1.208% |
| 69 | F | 1,731,958 | 22,639 | 1.307% |
| 70 | F | 1,674,584 | 24,422 | 1.458% |
| 71 | F | 1,647,705 | 26,607 | 1.615% |
| 72 | F | 1,707,812 | 29,424 | 1.723% |
| 73 | F | 1,259,662 | 25,287 | 2.007% |
| 74 | F | 1,238,863 | 26,098 | 2.107% |
| 75 | F | 1,199,896 | 28,571 | 2.381% |
| 76 | F | 1,218,702 | 31,604 | 2.593% |
| 77 | F | 1,056,331 | 30,493 | 2.887% |
| 78 | F | 960,647 | 30,611 | 3.186% |
| 79 | F | 898,590 | 31,718 | 3.530% |
| 80 | F | 831,640 | 33,085 | 3.978% |
| 81 | F | 788,013 | 34,649 | 4.397% |
| 82 | F | 715,575 | 35,705 | 4.990% |
| 83 | F | 671,025 | 37,703 | 5.619% |
| 84 | F | 631,230 | 38,788 | 6.145% |
| 85+ | F | 4,228,470 | 535,581 | 12.666% |

**Exhibit A9. Moratlity Load Development**

| | A | B | C | D = B/C | E = D/Yellow Cell |
|---|---|---|---|---|---|
| **Ages 0-64** | **Beneficiaries Observed in 2020 Data** | **Deaths Observed in 2020 Data** | **Deaths in population, assuming mortality follows that of a standard mortality table** | **Ratio of Deaths in Data to Deaths under Standard Mortality** | **Ratio / Denom Ratio (Covid Adjustment)** |
| Denominator | 2,891,098 | 88,961 | 16,802 | 5.295 | 1.000 |
| T1DM | 57,146 | 5,084 | 323 | 15.740 | 2.973 |

| **Ages 65+** | **Beneficiaries Observed in 2020 Data** | **Deaths Observed in 2020 Data** | **Deaths in population, assuming mortality follows that of a standard mortality table** | **Ratio of Deaths in Data to Deaths under Standard Mortality** | **Ratio / Denom Ratio (Covid Adjustment)** |
|---|---|---|---|---|---|
| Denominator | 17,716,452 | 904,081 | 727,555 | 1.243 | 1.000 |
| T1DM | 121,554 | 13,512 | 4,356 | 3.102 | 2.496 |

**Exhibit A10. CDC WONDER Demographic Projections**

CDC WONDER provides ten-year population projections from 2020 to 2030. The sample data below shows the projections by age, sex, ethnicity, and race.
We used the age, sex, and race specific projections to support our modeling projections over the 10 year period.
http://wonder.cdc.gov/wonder/help/PopulationProjections.html

| Age | Age Code | Gender | Gender Code | Ethnicity | Ethnicity Code | Race | Race Code | Year | Year Code | Projected Populations |
|---|---|---|---|---|---|---|---|---|---|---|
| < 1 year | 0 | Female | F | Hispanic | 2135-2 | American Indian or Alaska Native | 1002-5 | 2020 | 2020 | 14,408 |
| < 1 year | 0 | Female | F | Hispanic | 2135-2 | American Indian or Alaska Native | 1002-5 | 2030 | 2030 | 15,506 |
| < 1 year | 0 | Female | F | Hispanic | 2135-2 | Asian | 2028-9 | 2020 | 2020 | 5,172 |
| < 1 year | 0 | Female | F | Hispanic | 2135-2 | Asian | 2028-9 | 2030 | 2030 | 5,871 |
| < 1 year | 0 | Female | F | Hispanic | 2135-2 | Black or African American | 2054-5 | 2020 | 2020 | 28,203 |
| < 1 year | 0 | Female | F | Hispanic | 2135-2 | Black or African American | 2054-5 | 2030 | 2030 | 31,226 |
| < 1 year | 0 | Female | F | Hispanic | 2135-2 | Native Hawaiian or Pacific Islander | 2076-8 | 2020 | 2020 | 1,497 |
| < 1 year | 0 | Female | F | Hispanic | 2135-2 | Native Hawaiian or Pacific Islander | 2076-8 | 2030 | 2030 | 1,488 |
| < 1 year | 0 | Female | F | Hispanic | 2135-2 | White | 2106-3 | 2020 | 2020 | 449,143 |
| < 1 year | 0 | Female | F | Hispanic | 2135-2 | White | 2106-3 | 2030 | 2030 | 507,499 |
| < 1 year | 0 | Female | F | Hispanic | 2135-2 | Two or More Races | MULTI | 2020 | 2020 | 27,380 |
| < 1 year | 0 | Female | F | Hispanic | 2135-2 | Two or More Races | MULTI | 2030 | 2030 | 35,185 |
| | | | | | | ... | | | | |
| 100+ years | 100+ | Male | M | Not Hispanic | 2186-5 | American Indian or Alaska Native | 1002-5 | 2020 | 2020 | 92 |
| 100+ years | 100+ | Male | M | Not Hispanic | 2186-5 | American Indian or Alaska Native | 1002-5 | 2030 | 2030 | 193 |
| 100+ years | 100+ | Male | M | Not Hispanic | 2186-5 | Asian | 2028-9 | 2020 | 2020 | 683 |
| 100+ years | 100+ | Male | M | Not Hispanic | 2186-5 | Asian | 2028-9 | 2030 | 2030 | 1,070 |
| 100+ years | 100+ | Male | M | Not Hispanic | 2186-5 | Black or African American | 2054-5 | 2020 | 2020 | 1,953 |
| 100+ years | 100+ | Male | M | Not Hispanic | 2186-5 | Black or African American | 2054-5 | 2030 | 2030 | 3,419 |
| 100+ years | 100+ | Male | M | Not Hispanic | 2186-5 | Native Hawaiian or Pacific Islander | 2076-8 | 2020 | 2020 | 11 |
| 100+ years | 100+ | Male | M | Not Hispanic | 2186-5 | Native Hawaiian or Pacific Islander | 2076-8 | 2030 | 2030 | 22 |
| 100+ years | 100+ | Male | M | Not Hispanic | 2186-5 | White | 2106-3 | 2020 | 2020 | 15,099 |
| 100+ years | 100+ | Male | M | Not Hispanic | 2186-5 | White | 2106-3 | 2030 | 2030 | 26,437 |
| 100+ years | 100+ | Male | M | Not Hispanic | 2186-5 | Two or More Races | MULTI | 2020 | 2020 | 94 |
| 100+ years | 100+ | Male | M | Not Hispanic | 2186-5 | Two or More Races | MULTI | 2030 | 2030 | 177 |

**Exhibit A11. CDC WONDER Geographic Projections**

CDC WONDER provides ten-year population projections from 2020 to 2030. The sample data below shows the projections by five-year age-band sex, and state. We mapped regions on to the states, and then used the age, sex, and region specific projections to support our modeling projections over the ten-year period.
http://wonder.cdc.gov/wonder/help/PopulationProjections.html

| Age Group | Age Group Code | Gender | Gender Code | State | State Code | Year | Year Code | Projected Populations | Region |
|---|---|---|---|---|---|---|---|---|---|
| 0-4 years | 0-4 | Female | F | Alabama | 1 | 2020 | 2020 | 144,267 | South |
| 0-4 years | 0-4 | Female | F | Alabama | 1 | 2030 | 2030 | 152,260 | South |
| 0-4 years | 0-4 | Female | F | Alaska | 2 | 2020 | 2020 | 32,447 | West |
| 0-4 years | 0-4 | Female | F | Alaska | 2 | 2030 | 2030 | 36,024 | West |
| 0-4 years | 0-4 | Female | F | Arizona | 4 | 2020 | 2020 | 306,193 | West |
| 0-4 years | 0-4 | Female | F | Arizona | 4 | 2030 | 2030 | 369,980 | West |
| 0-4 years | 0-4 | Female | F | Arkansas | 5 | 2020 | 2020 | 98,414 | South |
| 0-4 years | 0-4 | Female | F | Arkansas | 5 | 2030 | 2030 | 107,398 | South |
| 0-4 years | 0-4 | Female | F | California | 6 | 2020 | 2020 | 1,534,935 | West |
| | | | | … | | | | | |
| 85+ years | 85+ | Male | M | Virginia | 51 | 2030 | 2030 | 84,778 | South |
| 85+ years | 85+ | Male | M | Washington | 53 | 2020 | 2020 | 49,304 | West |
| 85+ years | 85+ | Male | M | Washington | 53 | 2030 | 2030 | 74,169 | West |
| 85+ years | 85+ | Male | M | West Virginia | 54 | 2020 | 2020 | 14,486 | South |
| 85+ years | 85+ | Male | M | West Virginia | 54 | 2030 | 2030 | 18,683 | South |
| 85+ years | 85+ | Male | M | Wisconsin | 55 | 2020 | 2020 | 46,186 | Midwest |
| 85+ years | 85+ | Male | M | Wisconsin | 55 | 2030 | 2030 | 60,690 | Midwest |
| 85+ years | 85+ | Male | M | Wyoming | 56 | 2020 | 2020 | 3,946 | West |
| 85+ years | 85+ | Male | M | Wyoming | 56 | 2030 | 2030 | 5,867 | West |

**Exhibit A12. T1DM Mortality Calculations**

| Sample Calculation for a 45 Year old Male | | |
|---|---|---|
| **Metric** | **Value** | **Notes** |
| Standard Mortality Rate | 0.332% | From Exhibit A9 for a 45 year old Male |
| T1DM Mortality Load | 2.973 | Load for 0-64 year olds |
| T1DM Mortality Rate | 0.987% | Standard Mortality Rate * T1DM Mortality Load |

| Sample Mortality Reduction Calculation for a 45 Year old Male on CGMs and Insulin Pumps | | |
|---|---|---|
| **Metric** | **Value** | **Notes** |
| Reduction Factor | 50% | Reducation for CGMs and Pumps |
| T1DM Mortality Rate | 0.987% | Calculated above |
| Standard Mortality | 0.332% | Calculated above |
| T1DM mortality rate in excess of the standard mortality rate | 0.655% | T1DM Mortality Rate - Standard Mortality Rate |
| Reduction in Mortality Rate | 0.327% | Reduction Factor * T1DM mortality rate in excess of standard mortality rate |
| **New Mortality Rate** | **0.659%** | T1DM Mortality Rate - Reduction in Mortality Rate |
